# Virtual Primary Care for People with Opioid Use Disorder: A Scoping Review of Current Strategies, Benefits, and Challenges

**DOI:** 10.1101/2023.10.06.23296679

**Authors:** Shawna Narayan, Ellie Gooderham, Sarah Spencer, Rita McCracken, Lindsay Hedden

**Author notes:** **Correspondence to** Dr Lindsay Hedden, Faculty of Health Sciences, Simon Fraser University.

## Abstract

**Background:** There is a pressing need to understand the implications of the rapid adoption of virtual primary care for people with opioid use disorder. Potential impacts, including disruptions to opiate agonist therapies, and the prospect of improved service accessibility remain underexplored. This scoping review synthesizes current literature on virtual primary care for people with opioid use disorder, with a specific focus on benefits, challenges, and strategies.

**Methods:** We followed the Joanna Briggs Institute methodological approach for scoping reviews and reported our findings consistent with the Preferred Reporting Items for Systematic reviews and Meta-Analyses extension for Scoping Reviews. We conducted searches on MEDLINE, Web of Science, CINAHL Complete, and Embase using our developed search strategy with no date restrictions. We incorporated all study types that included the three concepts (i.e., virtual care; primary care; people with opioid use disorder). We excluded research on minors, asynchronous virtual modalities, and care not provided in a primary care setting. We used Covidence to screen and extract data, pulling information on study characteristics, health system features, patient outcomes, and challenges and benefits of virtual primary care. We conducted inductive content analysis and calculated descriptive statistics. We appraised the quality of studies using the Quality Assessment with Diverse Studies tool and categorized findings using the Consolidated Framework for Implementation Science.

**Results:** Our search identified 1474 studies. We removed 536 duplicates, leaving 936 studies for title and abstract screening. After a double review process, we retained 28 studies for extraction. Most studies described virtual primary care delivered via phone (n=18, 64.3%) rather than video. While increased healthcare accessibility was a significant benefit (n=13, 46.4%) to the adoption of virtual visits, issues around access to technology and digital literacy stood out as the main challenge (n=12, 42.9%).

**Conclusions:** The available studies highlight the potential for enhancing accessibility and continuous access care for people with opioid use disorder using virtual modalities. Future research and policies must focus on bridging gaps to ensure virtual primary care does not exacerbate or entrench health inequities.

## INTRODUCTION

The opioid crisis remains a significant public health challenge [1], highlighted by the rising number of deaths globally [2–4]. In North America, illicit drug poisoning deaths can be attributed to a contaminated drug supply saturated with synthetic opioids, particularly fentanyl [5]. Simultaneously, opioid use disorder (OUD) is on the rise [6], with 21.4 million cases worldwide in 2019 [7]. Effective medications for opioid use disorder, such as opioid agonist therapies (OAT), have shown potential in reducing illicit drug poisoning deaths [8, 9]. Given that primary care services are often at the forefront of diagnosing and treating OUD, evidence shows that accessible primary care services can help achieve better health outcomes for people with opioid use disorder (PWOUD) [10, 11].

The COVID-19 pandemic introduced new barriers to traditional approaches to OUD treatment, including in-person primary care services, due to the need for social distancing and reduced in-person interactions [12, 13]. In response to COVID-19 public health measures, healthcare systems implemented remote care solutions, transforming the delivery of healthcare services [14] and accelerating the adoption and utilization of virtual healthcare [15–17]. During this same period, much of the progress that had been made to address the opioid crisis was reversed [18–20]; illicit drug poisoning deaths increased as a result of limited health services [21], poisoned supply [22], and solitary drug use [23]. In the context of these challenges, and as a result of pandemic-related public health measures, virtual care emerged as a potential solution to address the unique needs of PWOUD by providing essential healthcare services remotely [24].

The widespread shift to virtual care facilitated continuous access to care during the pandemic. This shift may have been helpful for individuals with OUD [25, 26], many of whom experience barriers to health care due to intersecting effects of criminalization, discrimination, and social marginalization [13]. Virtual care allows clinicians to deliver essential healthcare services to PWOUD [27, 28], offering improved accessibility [29] and the ability to deliver OAT [30], while reducing barriers related to transportation, mobility limitations, and stigma [31]. However, virtual visits may limit clinicians’ ability to fully assess patients’ physical and emotional wellbeing [27] and may introduce technological barriers for those with limited internet access and low digital literacy [32, 33]. Additionally, virtual care may not be suitable for all aspects of OUD treatment as some interventions, such as injectable OAT [34], require regular in-person management [27].

Given that virtual platforms have become increasingly common for primary care – a trend that is expected to persist beyond the pandemic [35] – understanding the benefits and limitations of virtual care interventions is essential for shaping future healthcare delivery models and optimizing the care provided to patients with social and clinical complexities. This scoping review aims to synthesize the current knowledge landscape, research gaps, and offer insights into the potential of virtual primary care for PWOUD.

## METHODS

### Overall study design

This review informs a mixed-methods study investigating changes to primary care access and patient outcomes following the rapid introduction of virtual care for PWOUD in British Columbia, Canada [36]. We conducted the review in accordance with the Joanna Briggs Institute Reviewer Manual [37, 38] and the framework suggested by Arksey and O’Malley [39]. We registered our protocol on the Open Science Framework [40] and report our findings using the Preferred Reporting Items for Systematic Reviews and Meta-Analyses extension for Scoping Reviews (PRISMA-ScR) [41] and the PRISMA for Abstracts Checklist [42] (see supplementary materials). Manuscript editing was supported through a large language model (i.e., ChatGPT [43]) to improve clarity and conciseness [44]. Authors reviewed generated text to ensure original meaning.

### Eligibility criteria

We established our eligibility criteria *a priori*. We included all study types (abstracts, viewpoints, observational studies, qualitative data, systematic reviews, etc.) with a focus on the three concepts (i.e., virtual care; primary care; PWOUD). We considered all studies that described the use of any form of synchronous virtual visits, such as telephone calls and video conferencing for the purpose of providing primary care services (i.e., prevention, screening, diagnosis, treatment) for PWOUD; this is not exclusive to providing OAT. We included adult populations with opioid use disorder and did not restrict based on the presence or absence of other comorbidities. No geographical restrictions were implemented; however, we restricted the search to studies published in English.

We excluded any study that did not include all three concepts (virtual care; primary care; PWOUD). We defined *primary care* as “the provision of integrated, accessible health care services by physicians and their health care teams who are accountable for addressing a large majority of personal health care needs, developing a sustained partnership with patients, and practicing in the context of family and community” [45]. Therefore, primary care settings are those where patients could receive comprehensive care (i.e., treatment for any type of health care issue) [46], including physician’s offices, community health centers, and specific outpatient clinics. We included some settings that may not traditionally be considered primary care – such as certain opioid treatment programs, syringe service programs, or veterans’ health association programs and clinics – when there was evidence that the setting also attended to health care issues beyond OUD [47–49]. We did exclude, however, psychiatric clinics, addiction clinics, some opioid treatment programs, and syringe services programs where these solely focused on treating OUD or did not report providing other comprehensive care. Other exclusions included studies taking place in prisons or hospitals; studies with participants under the age of 18; studies that described asynchronous virtual visits (e.g., text messaging, mobile applications); studies that did not specify the practice settings or the type of virtual modality. Additionally, studies that reported on substance use disorders but did not present data specific to OUD were excluded.

### Search Strategy

We developed search strategies [50–54] by combining MesH terms (e.g., Analgesics, Opioids; Telemedicine; Primary Health Care) and free-text keywords (e.g., Methadone, Virtual, ‘Family Doctor’) using Boolean (e.g., AND, OR, NOT), truncation and wildcard operators. Librarians from the University of British Columbia and Simon Fraser University verified the strategies. From 6 December 2022 to 8 December 2022, one reviewer (SN) systematically searched MEDLINE® (Ovid) [51], CINAHL Complete (EBSCOhost) [53], EMBASE (Ovid) [52], and the Web of Science Core Collection [50] with no end date. From May to June 2023, we searched grey literature using the Canada Commons database [54], Google Scholar, Trip, Greymatters, and select organization websites (i.e., Canada Health Infoway, BC Centre on Substance Use, Centre for Addiction and Mental Health, Canadian Research Initiative in Substance Misuse). Following the search and extraction, the bibliographies of included studies were systematically checked for additional references [55].

### Screening

One reviewer (SN) identified and removed all duplicates using Covidence [56], a systematic review software, after which two reviewers (SN and EG) screened the identified studies in a two-stage screening process (title/abstract screening and full-text screening). We achieved a 75% minimum agreement in a pilot test of our screening and reviewed any conflicts. During screening, some studies did not have well-defined care settings, leaving raters to apply inclusion criteria differently. Furthermore, the studies spanned a range of methodologies and outcomes, introducing additional challenges in achieving consistent ratings. We reconciled discrepancies at both stages through discussion with the research team and recorded the reasons for study exclusion at each step.

### Data extraction and analysis

We (EG, LH, RM and SN) created a data extraction chart (see supplementary materials) to capture relevant information (i.e., setting, benefits, challenges, virtual modality, patient population) in Covidence. To ensure its efficacy, we first piloted this chart on a small sample of studies. Based on insights from this pilot, we refined the chart to better capture the nuances of health service features specific to primary care settings. This revised chart became the cornerstone for the subsequent data extraction from the studies. In parallel, EG and SN conducted quality appraisals of the studies using the Quality Assessment with Diverse Studies (QuADS) tool [57]. No study that reached data extraction was excluded due to their QuADS score. Following data extraction and quality appraisal, SN and EG reviewed the data to reach a consensus. We transferred the harmonized data to Microsoft Excel to conduct inductive content analysis, which led us to identify patterns, which we subsequently organized into themes.

For theme categorization, we utilized the Consolidated Framework for Implementation Science (CFIR) [58], a widely recognized meta-theoretical model previously applied in other virtual care reviews [59–61]. The CFIR is comprised of five major domains (Figure 1):

1. Innovation: the change being implemented. In the context of our scoping review, we equate this with synchronous virtual visits in primary care.
2. Outer Setting: the context in which the Inner Setting exists. For this review, the outer setting is the state, including laws, policies, and institutions which inform the provision and influence access to primary care.
3. Inner Setting: the more immediate context where the innovation is applied. While virtual care was implemented throughout the healthcare system, we are specifically interested in how it has been applied and experienced in primary care.
4. Individuals: the actors involved in the provision and experience of the given innovation. Within the primary care milieu, there are two principal actors. ‘Innovation Receivers’ – in this study, PWOUD – and ‘Innovation Deliverers’ – clinicians offering virtual primary care.
5. Implementation Process: the methods and strategies employed to implement the innovation. This includes facilitators and barriers that influence implementation of virtual care.

**Figure 1.**
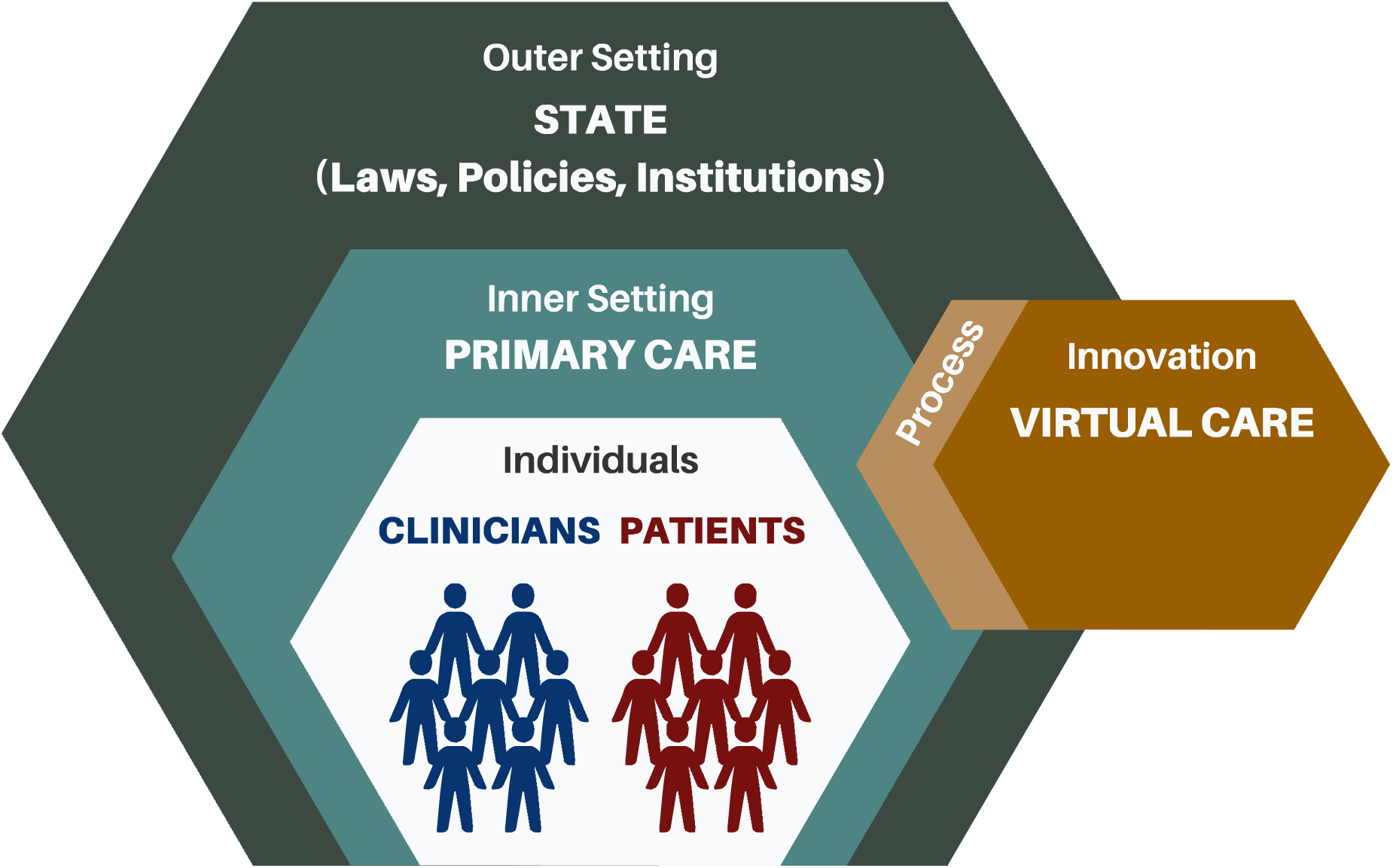
Consolidated Framework of Implementation Science Domains.

We discuss facilitators and barriers to the Implementation Process (Domain 5) within each of the other four domains.

## RESULTS

We identified a total of 1474 studies from our searches of Embase (OVID) (n=780), Web of Science Core Collection (n=283), Ovid MEDLINE (n=263), CINAHL Complete (EBSCOhost) (n=142) databases and through citation searching (n=6). After removing 536 duplicates, we title and abstract screened 936 studies, retaining 155 studies for full-text review. After a double-review process, we identified 28 papers for data extraction and quality assessment. A PRISMA flow diagram details the exclusions (Figure 2).

**Figure 2.**
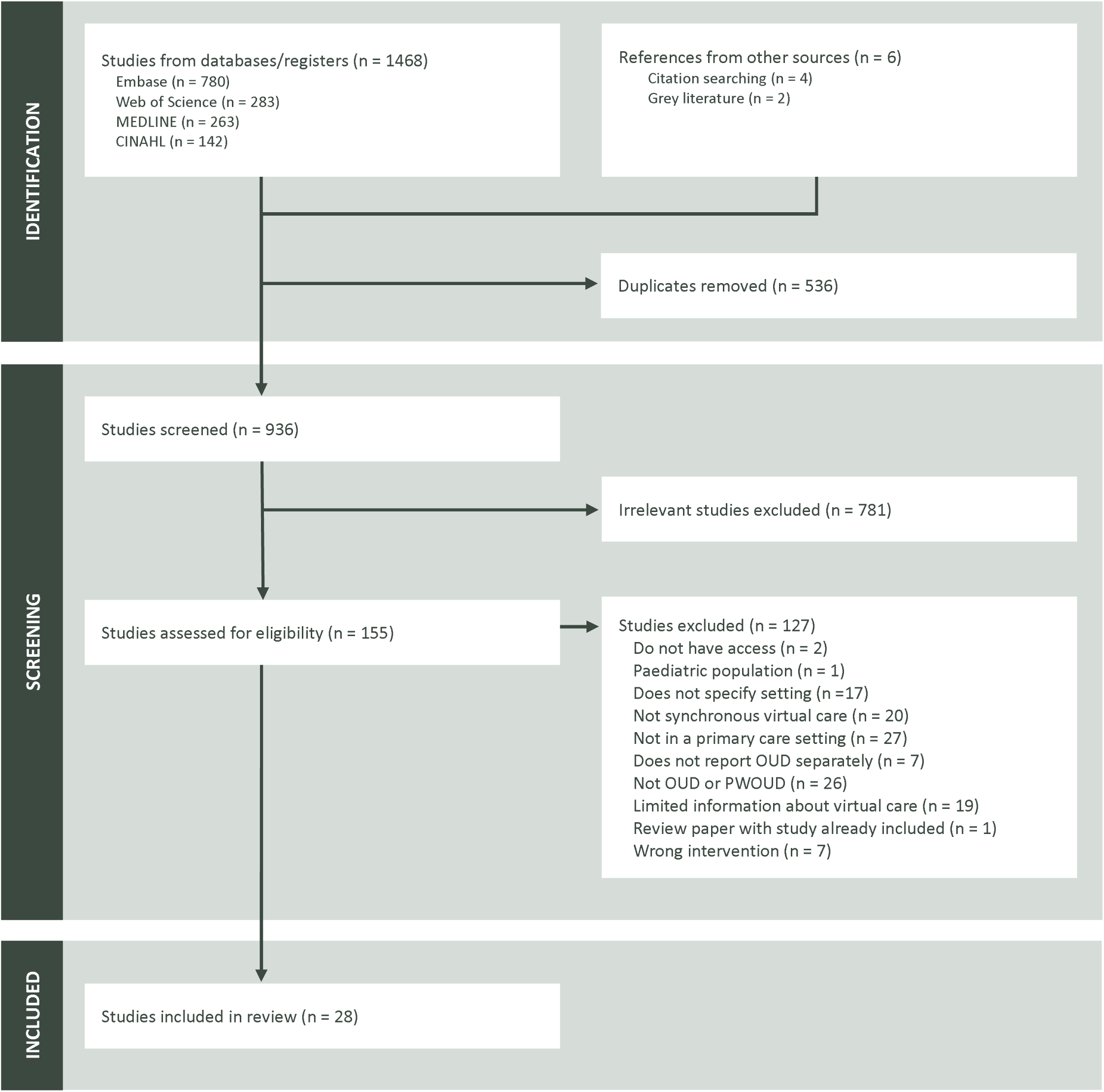
PRISMA diagram.

### Study Characteristics

The majority of studies were based in the United States and were published after the emergence of COVID-19 (Table 1). Only four studies were conducted prior to the pandemic [62–65], emphasizing the paucity of research on virtual primary care for PWOUD conducted before 2020. Original research constituted the largest group of studies (n=11, 39.3%), employing various study designs. Most studies represented either quantitative (n=15, 53.6%) or qualitative (n=4, 14.3%) designs, while one was mixed methods. Other studies reported findings as commentaries (n=7, 25.0%) or economic evaluations (n=1, 3.6%).

**Table 1.**
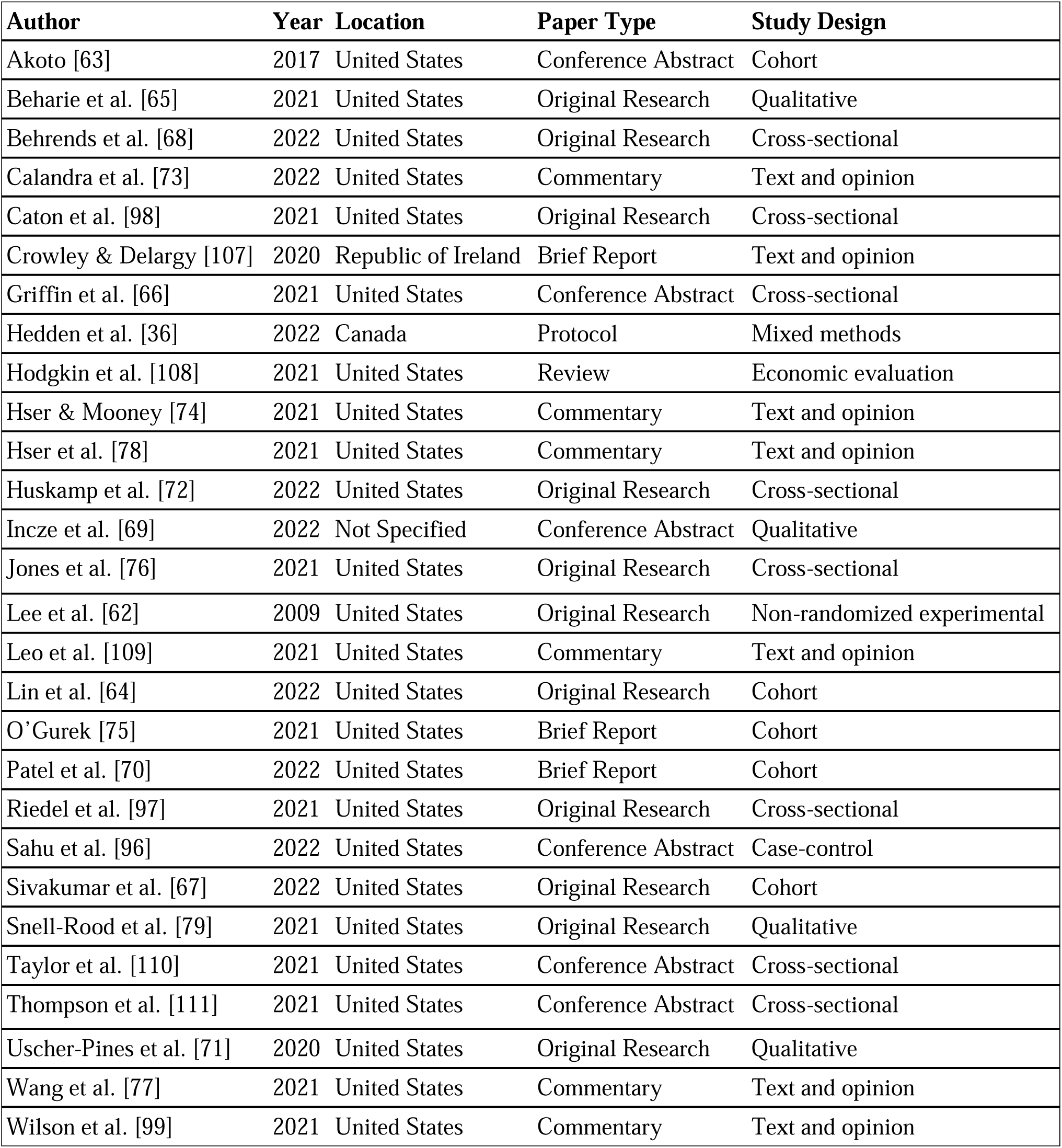
Study Characteristics.

The key objectives and results identified in each study are summarized in Table 2. Nineteen (67.9%) studies identified incorporating virtual care as part of their primary study purpose or design, while the remaining studies included virtual care as an ancillary component of their research (n=9, 32.1%). Studies commonly reported the introduction of virtual modalities due to COVID-19 (n=17, 60.7%), with a few of these papers also examining changes to or increases in existing virtual modality use in response to the pandemic (n=6, 21.4%). Outcomes included reports of shifts in health service utilization, such as changes to appointment modalities (n=16, 55.2%), frequency or demand (n=4, 14.3%), and differences in prescription duration (n=4, 14.3%). Four (14.3%) studies specify participants’ gender [65, 68, 76, 79], while four (14.3%) specify participants’ sex [64, 67, 70, 75]. Two (7.1%) studies do not clarify whether they are reporting sex or gender [62, 110], and the remaining papers (n=18, 64.3%) do not mention either category. Benefits and challenges of utilizing virtual primary care for PWOUD are grouped within the CFIR domains below and in Table 3.

**Table 2.**
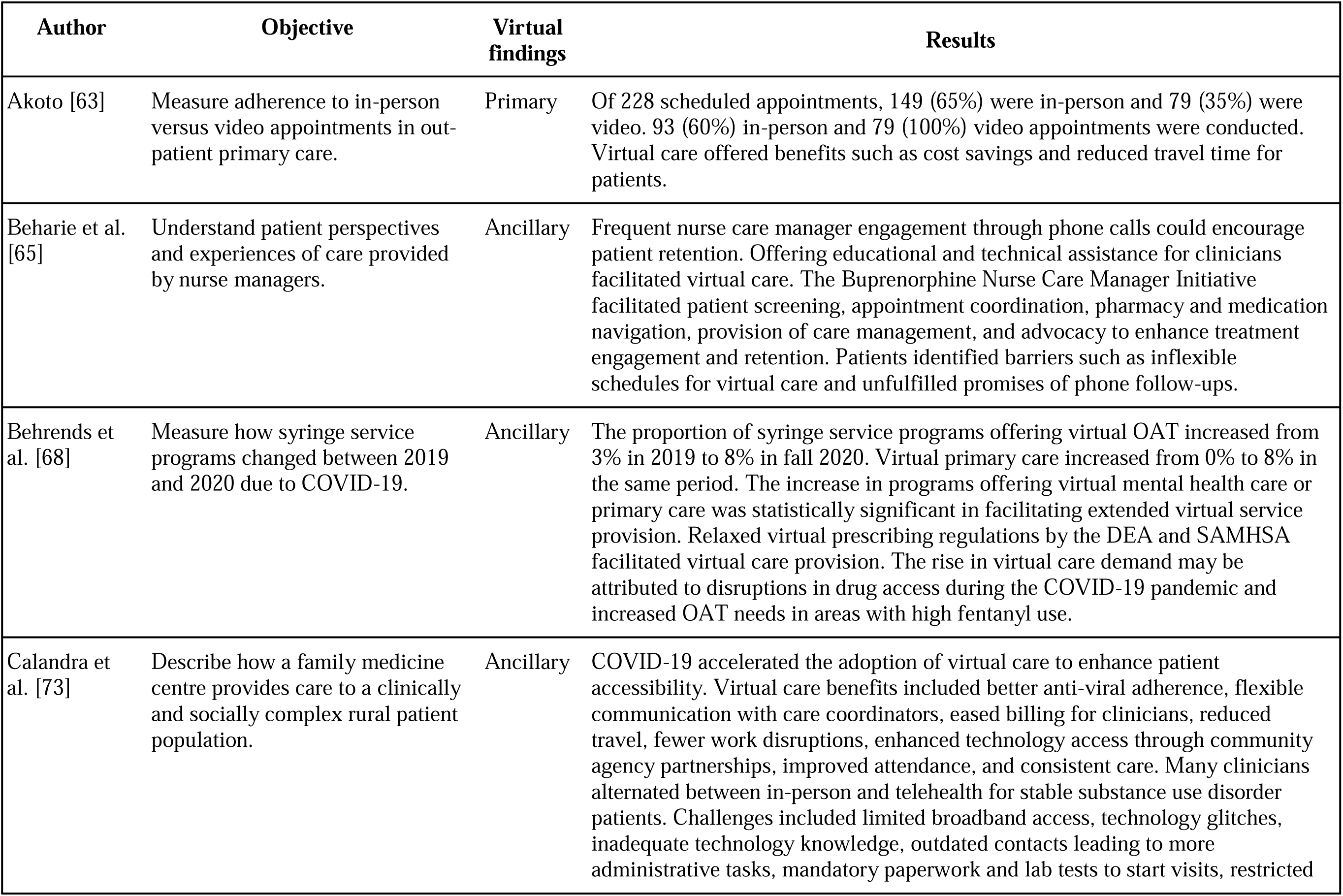
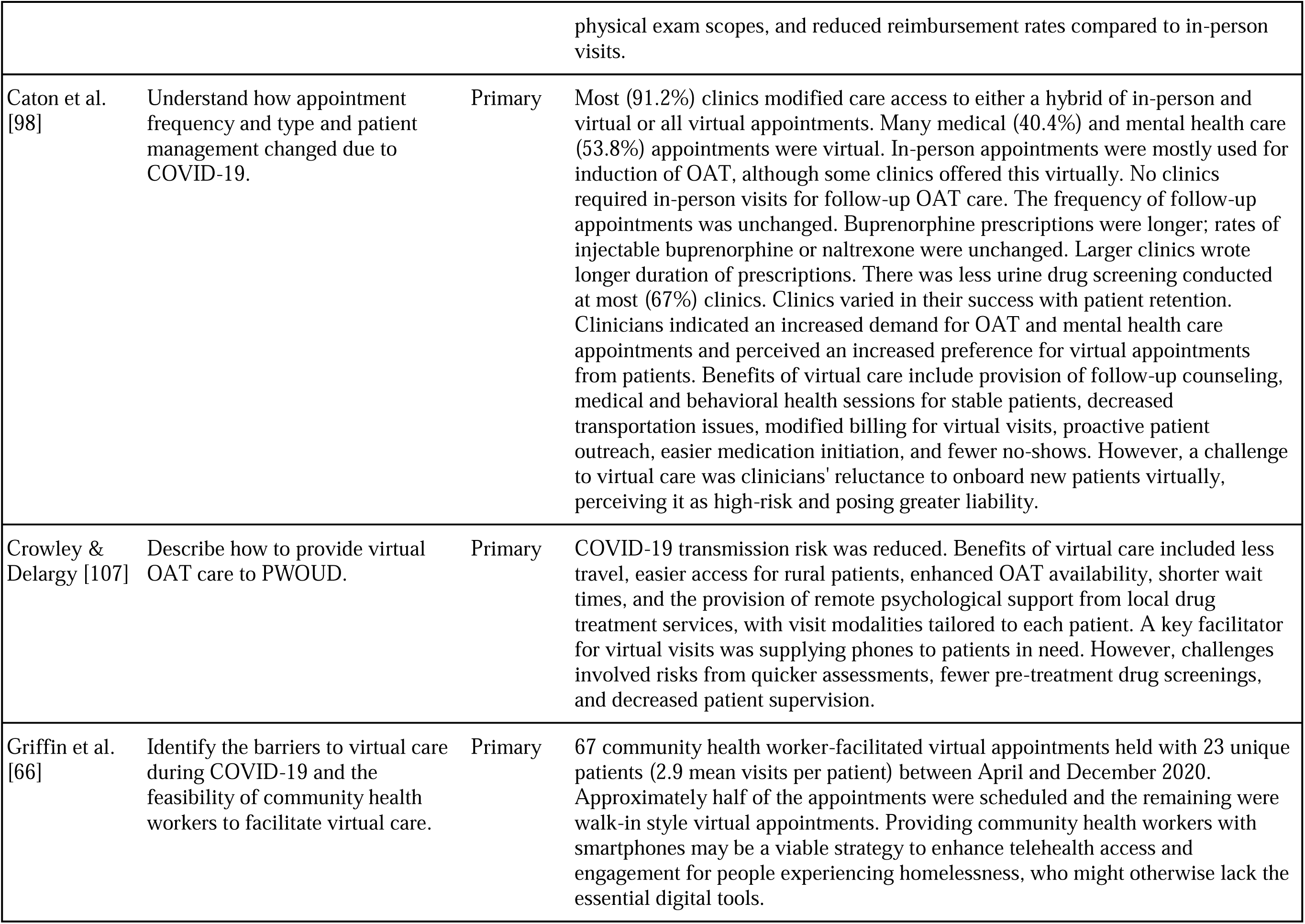
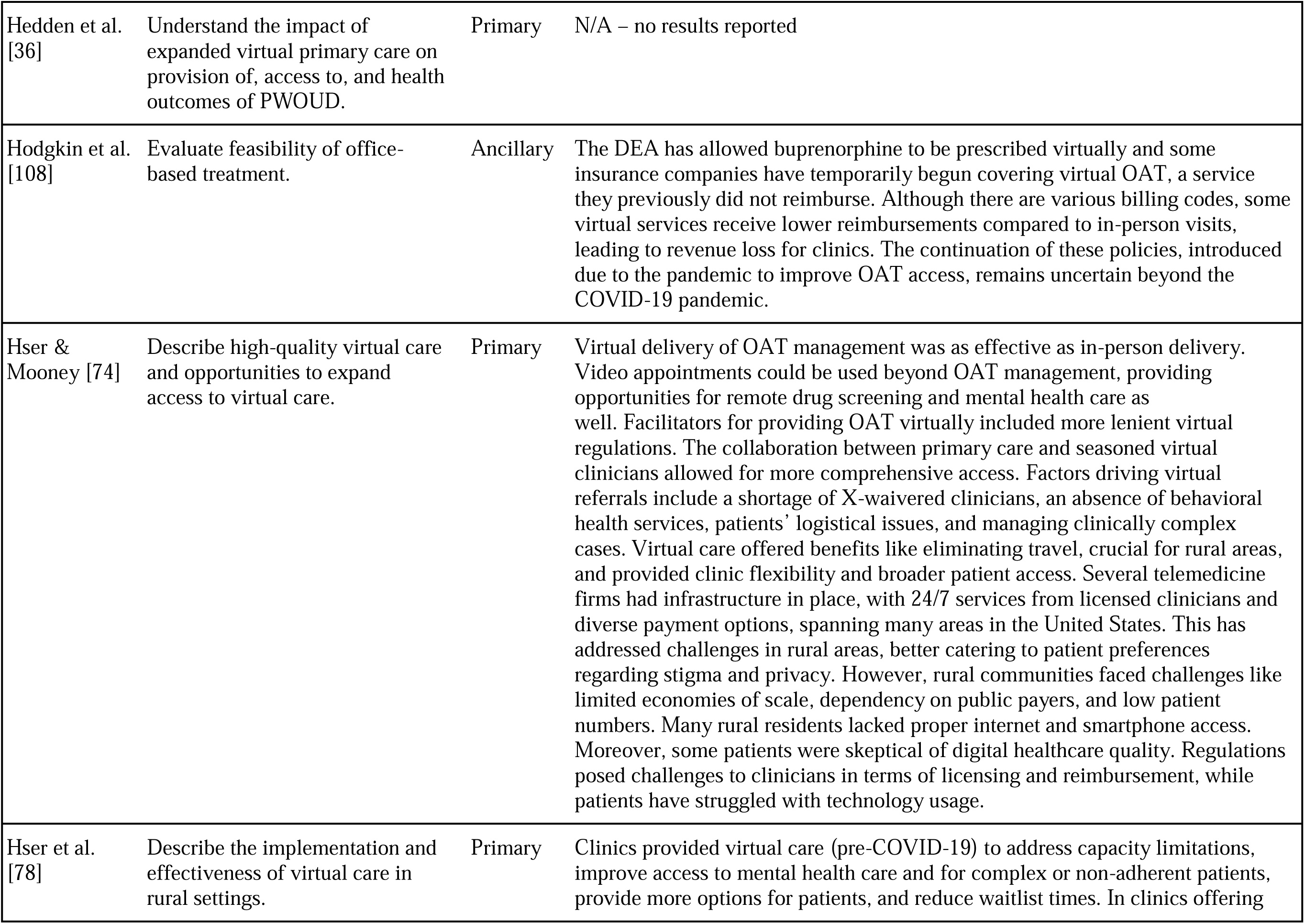
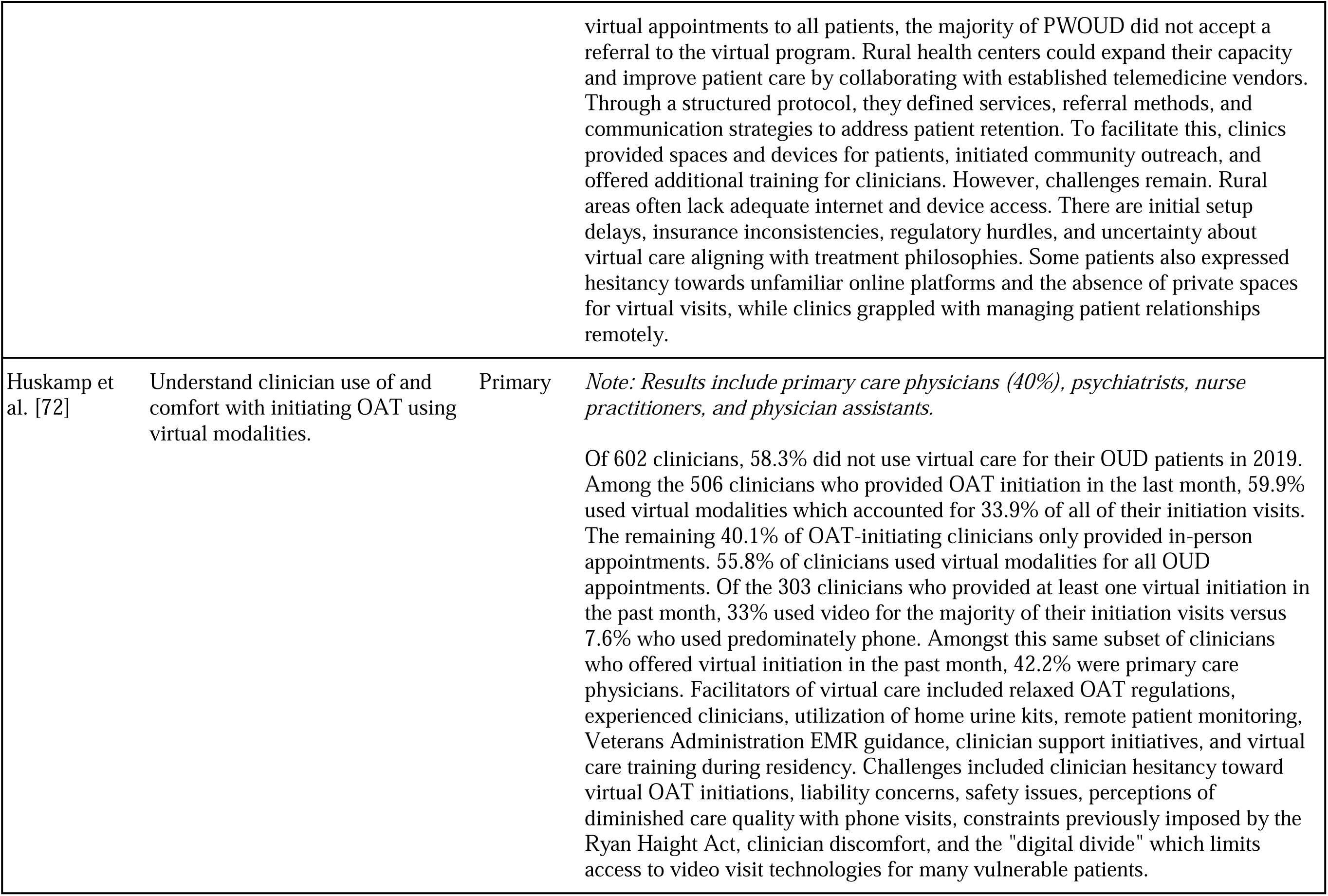
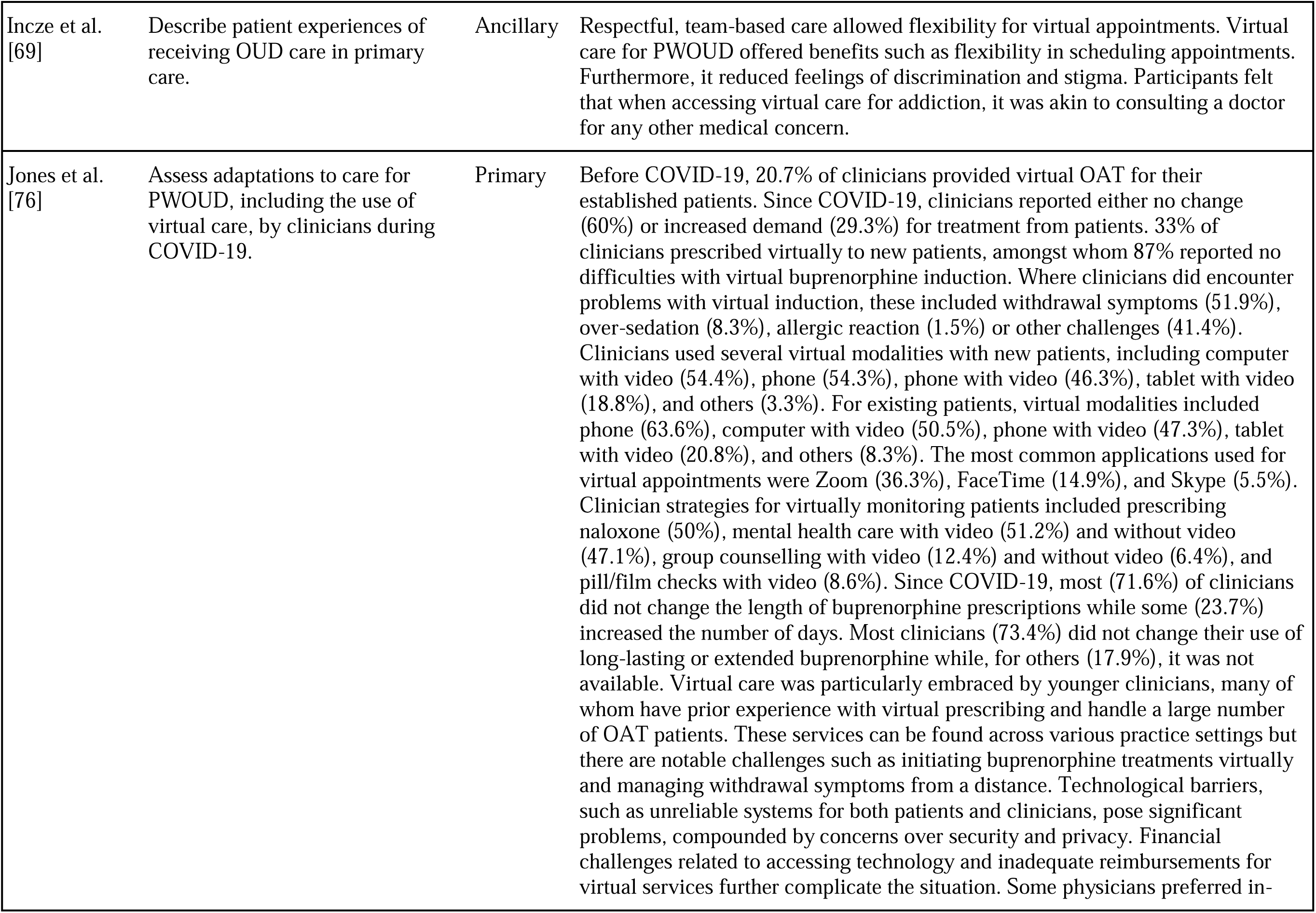
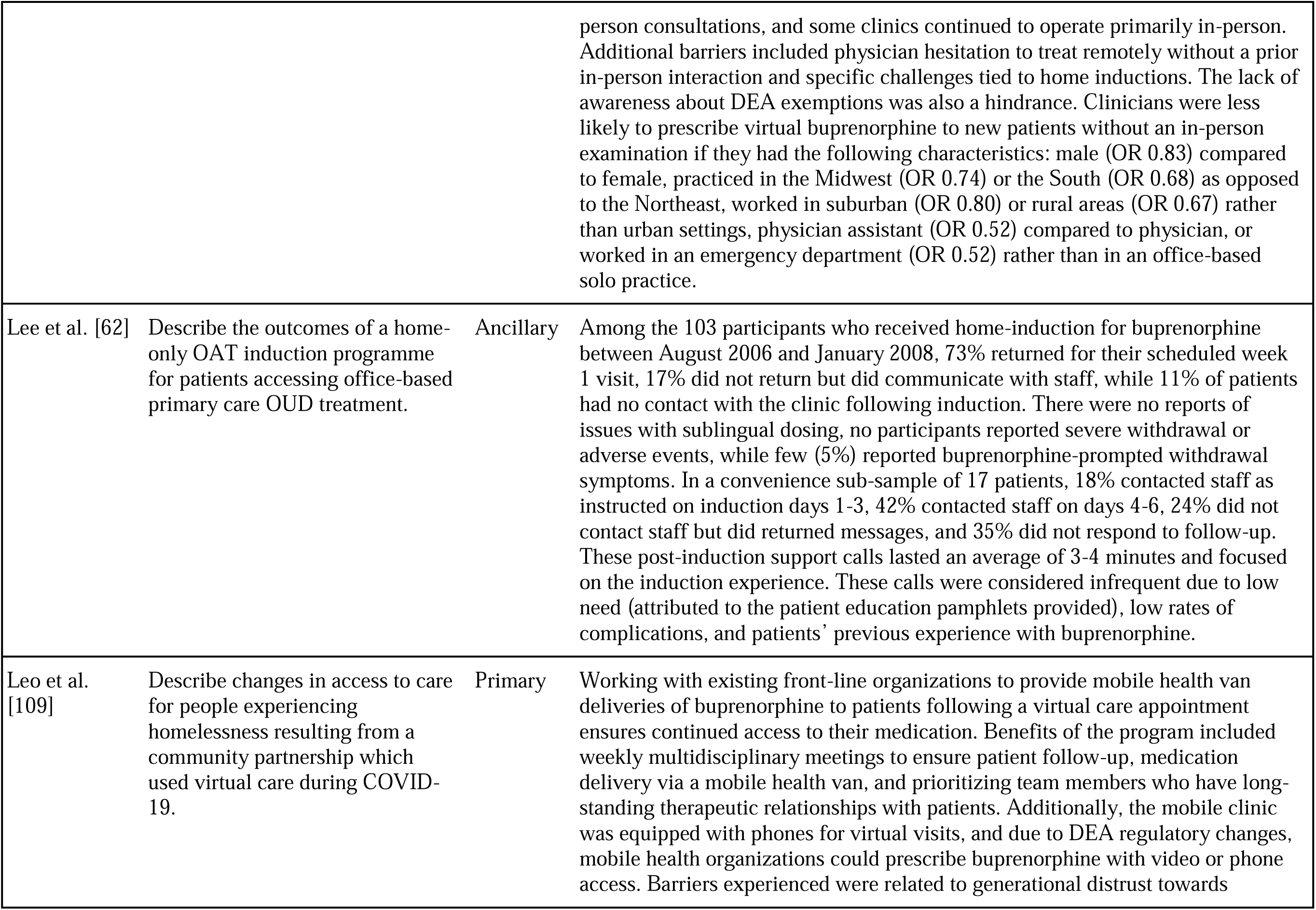
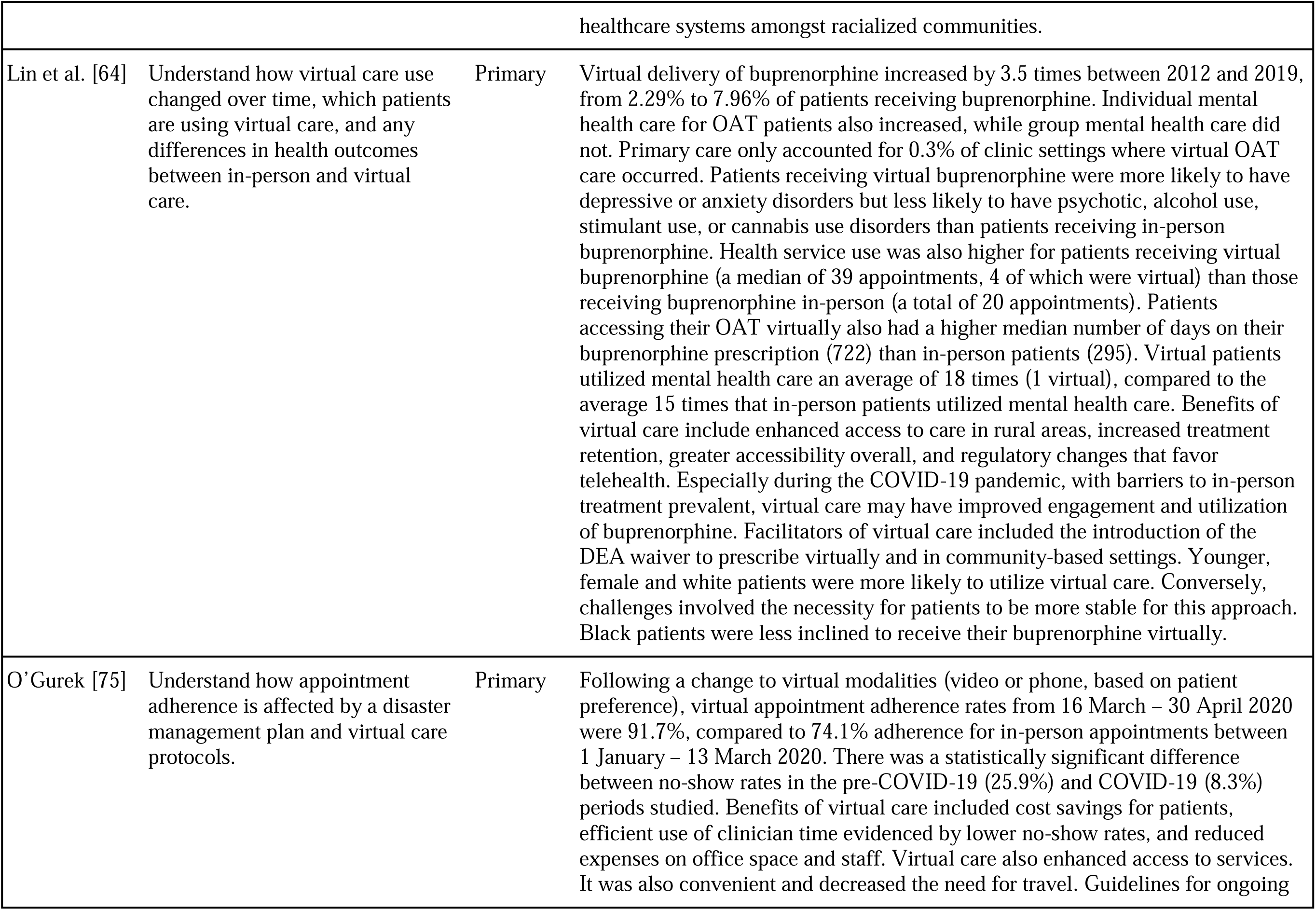
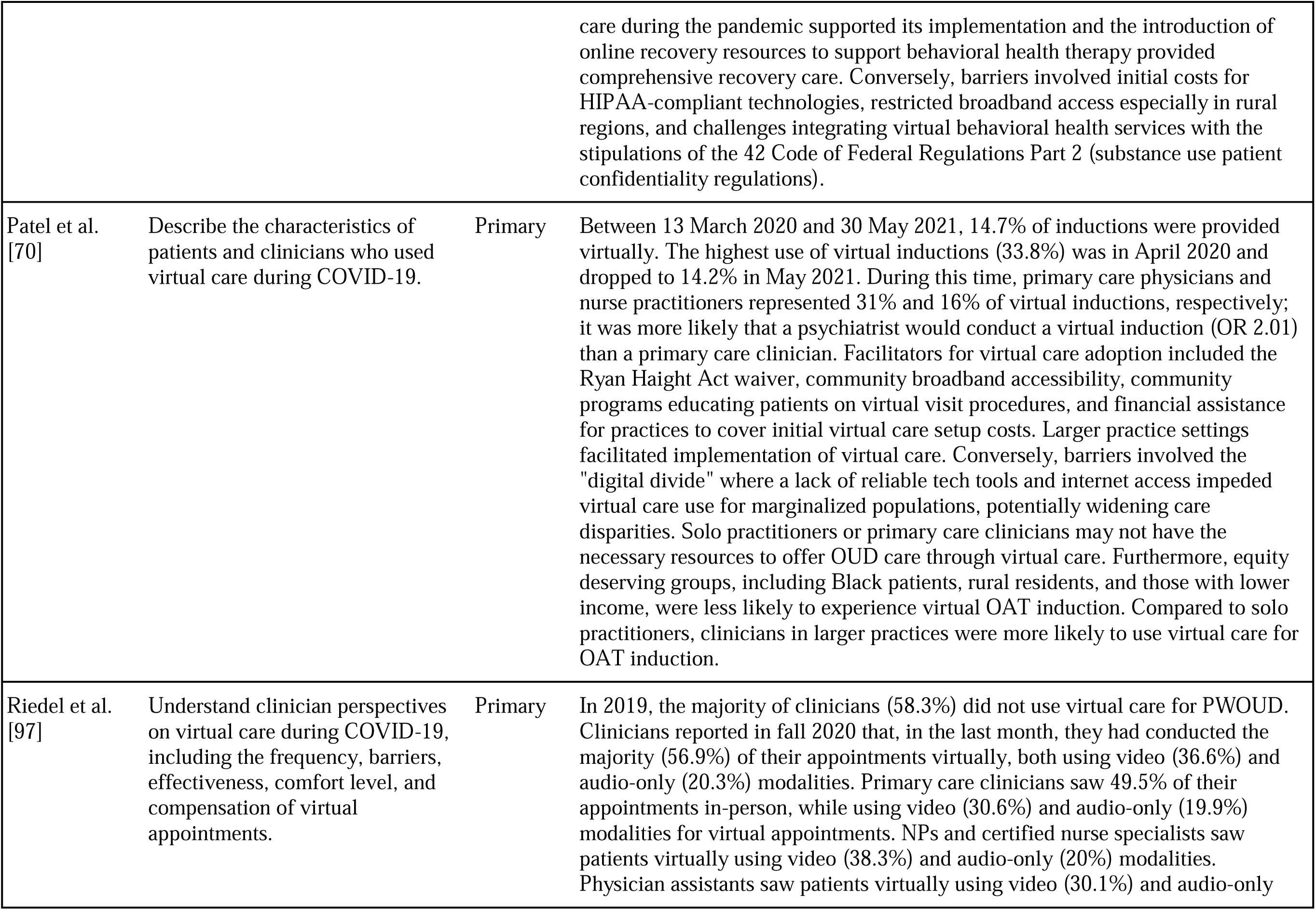
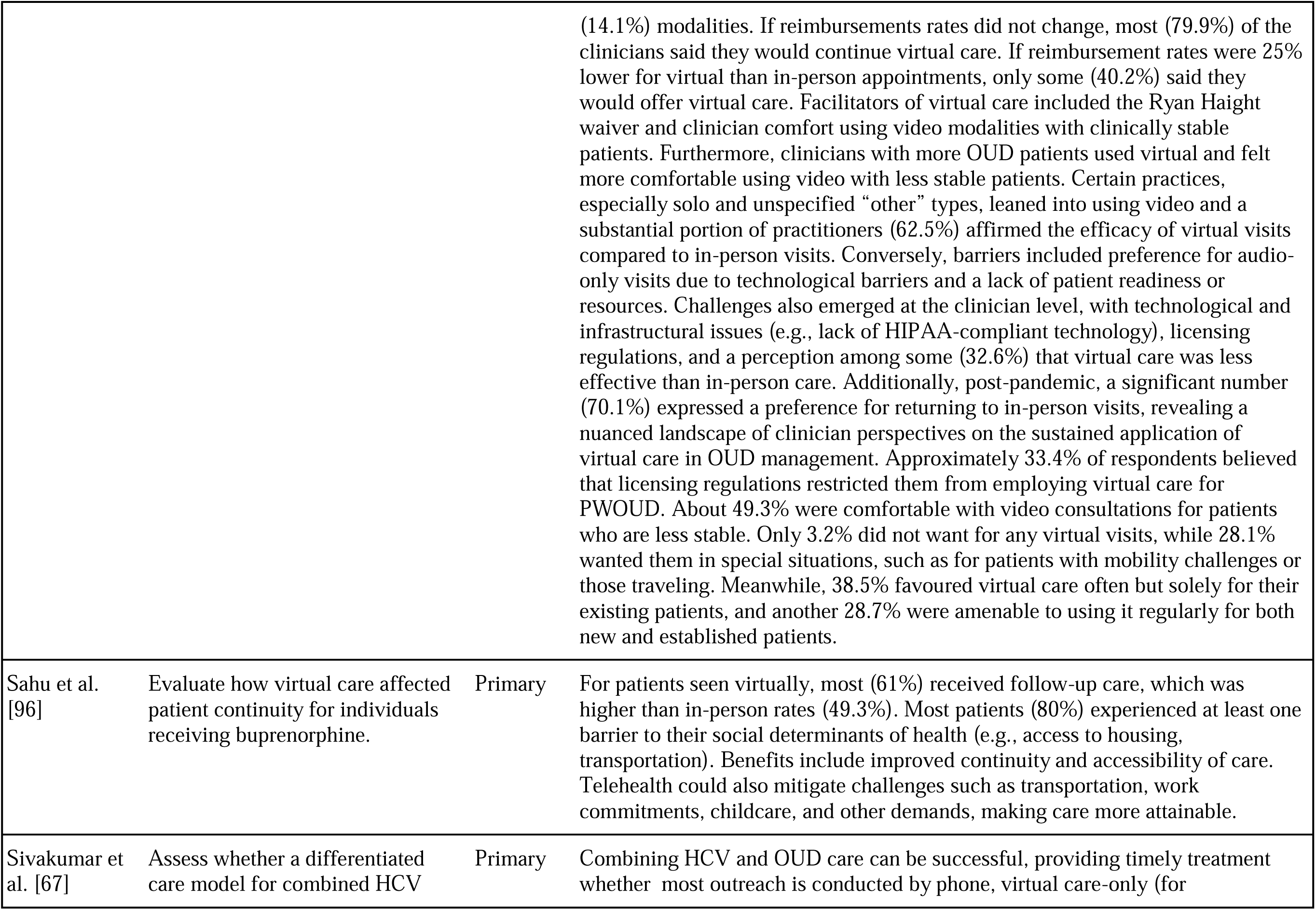
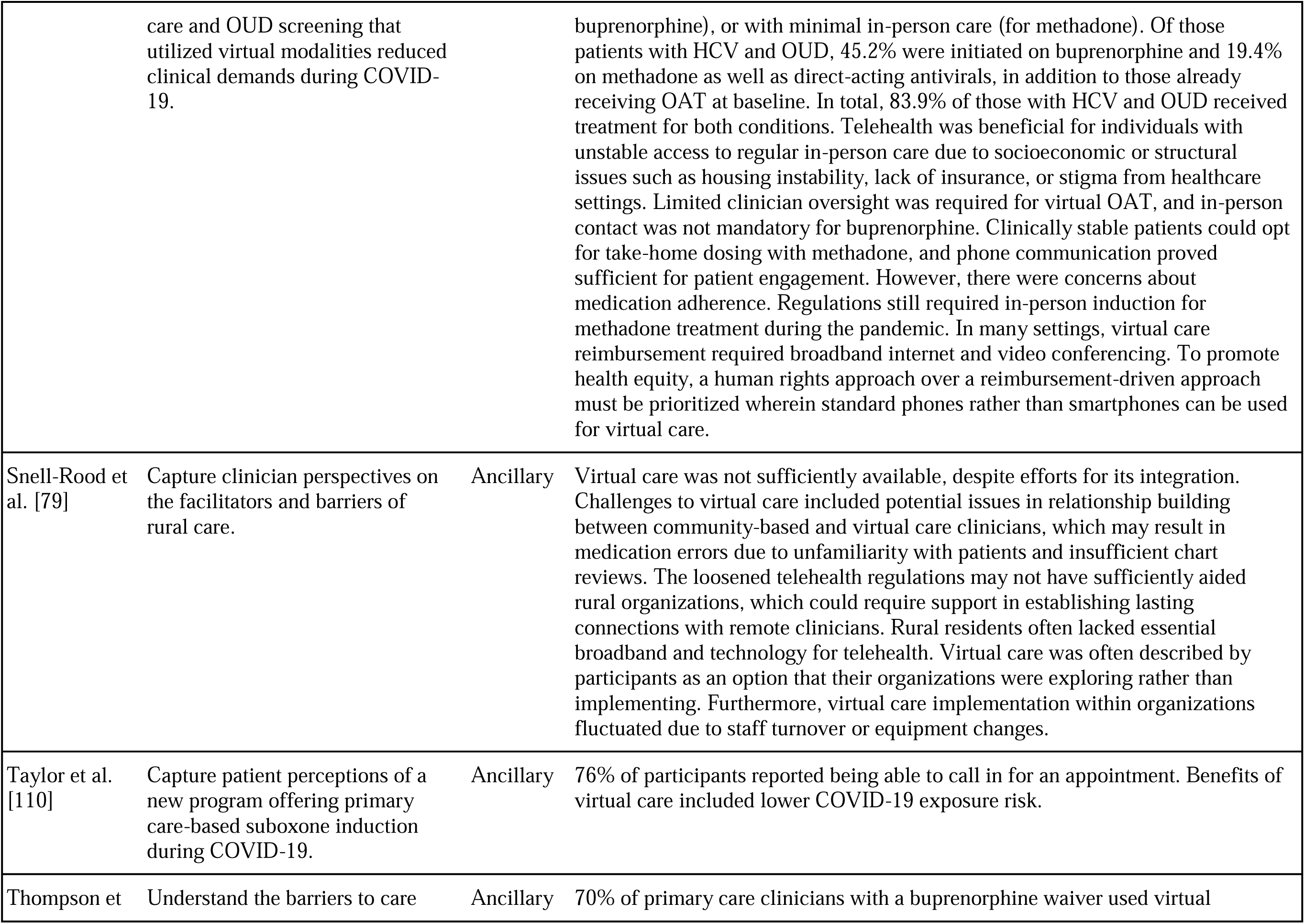
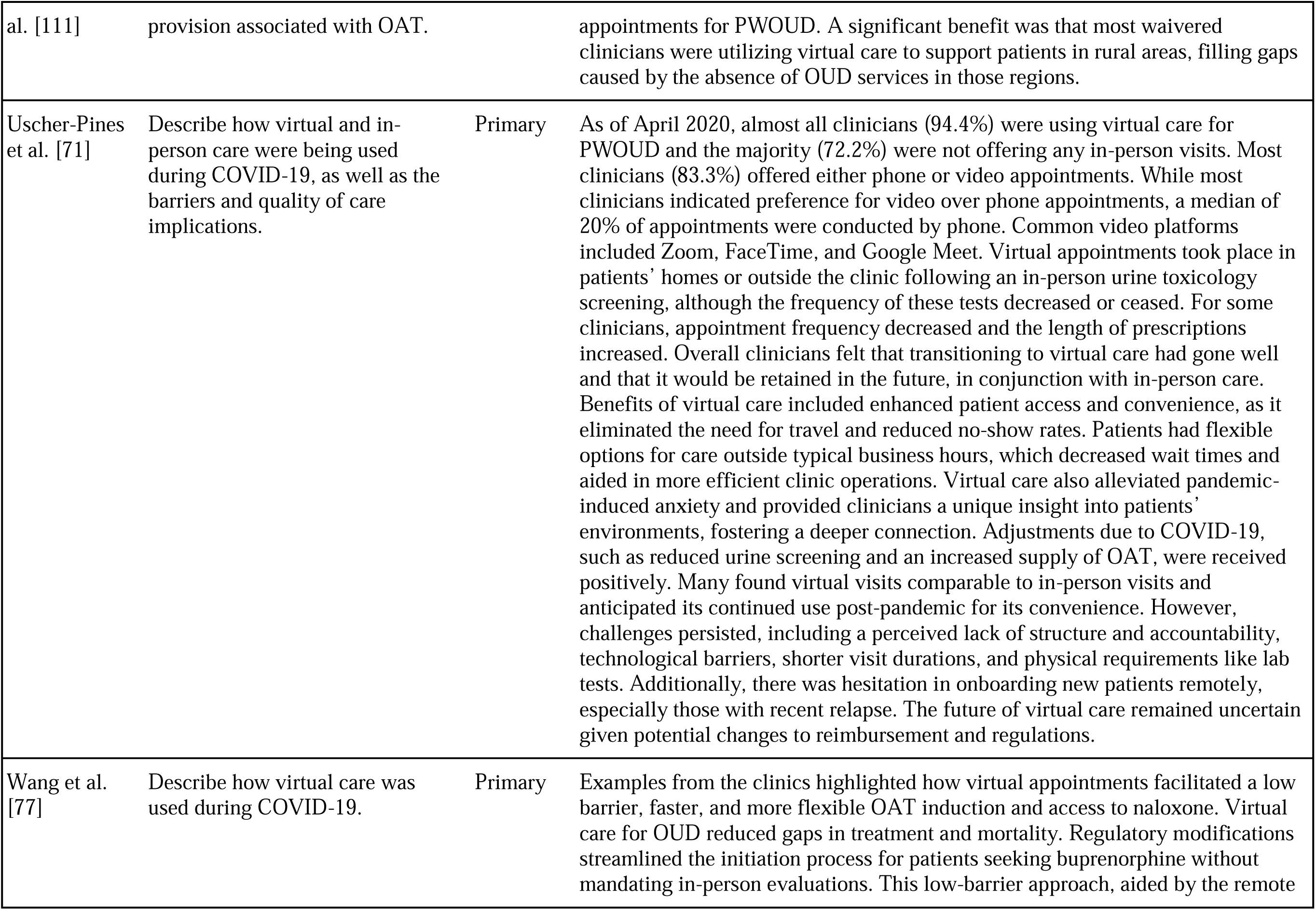
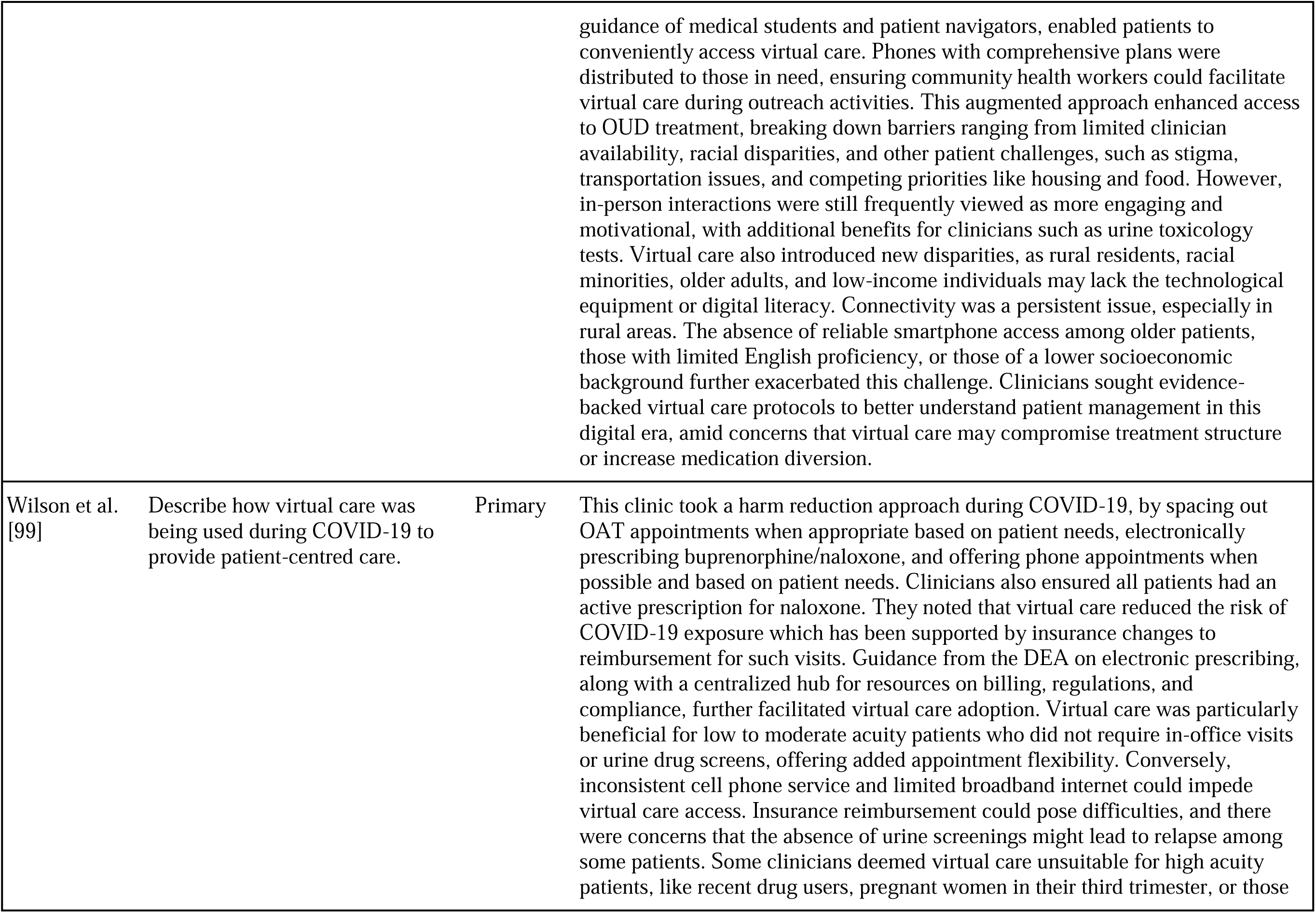
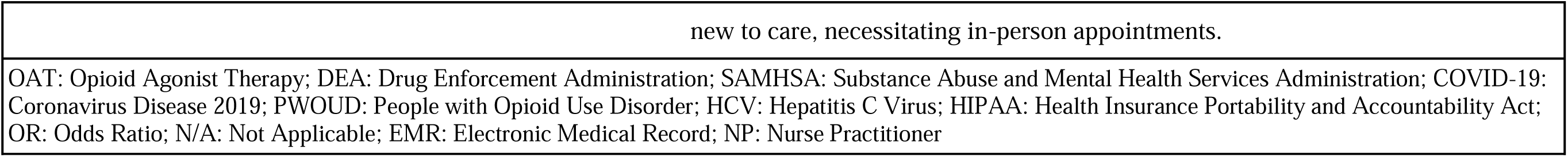
Study objectives and virtual primary care specific results.

**Table 3.**
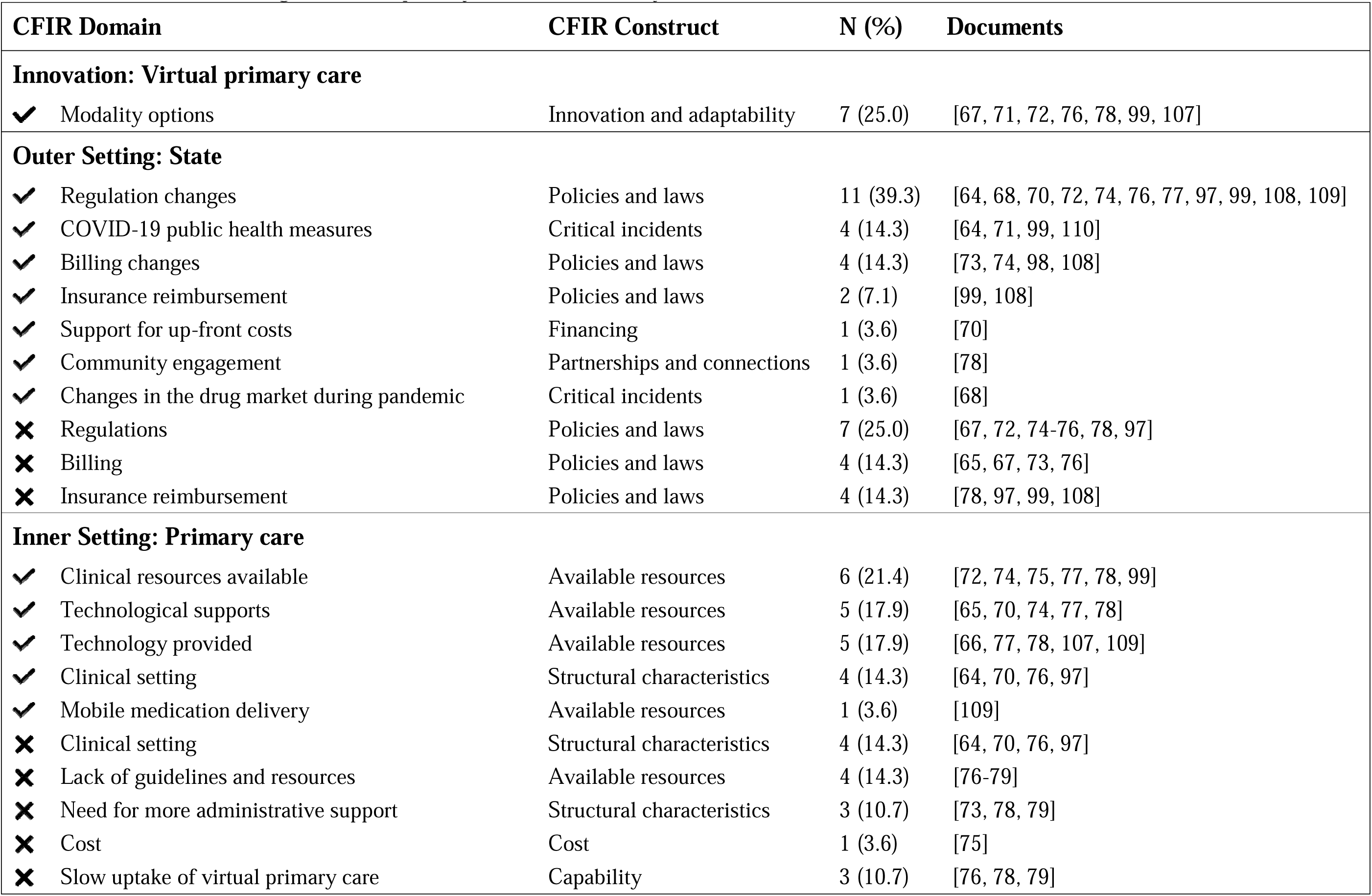
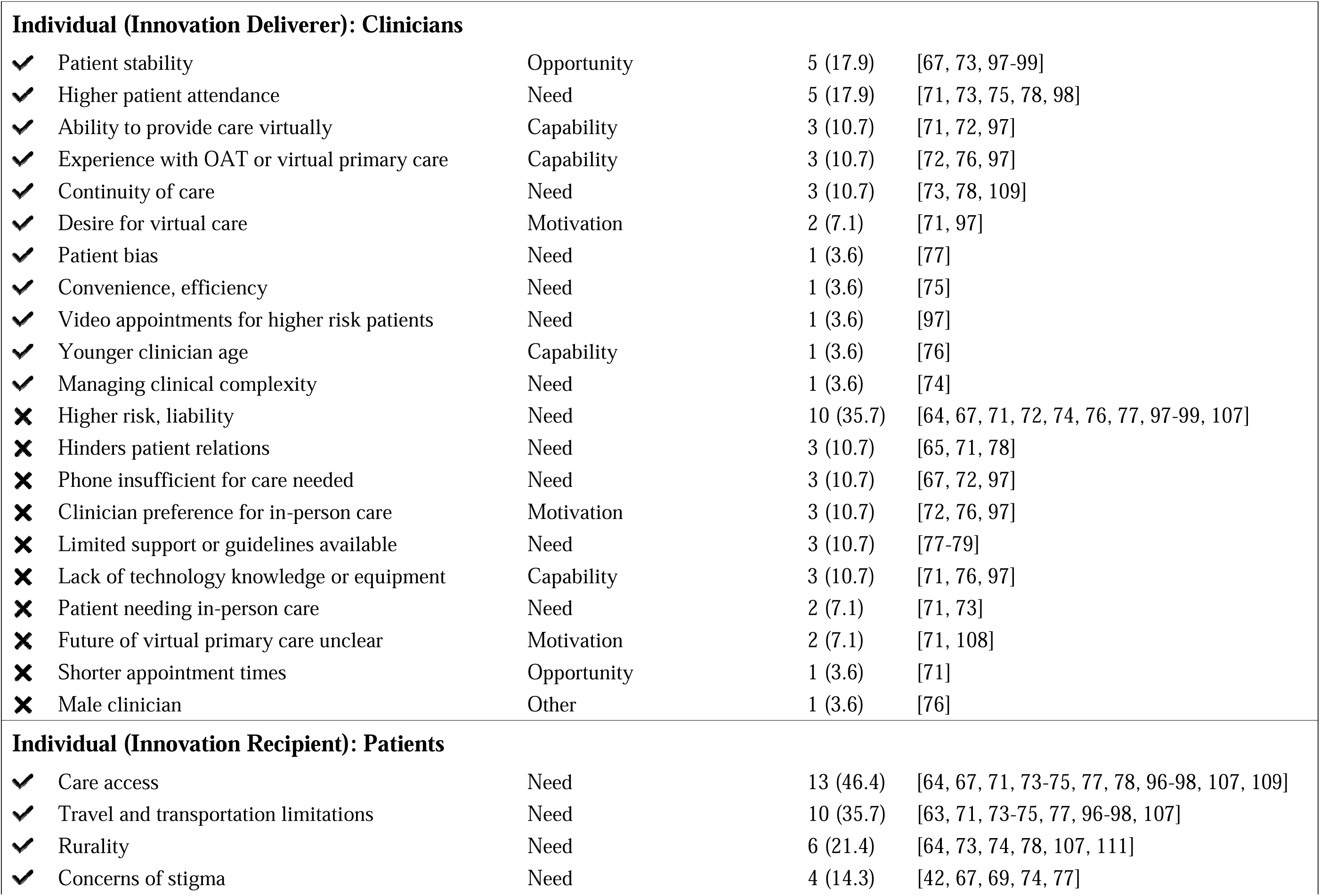
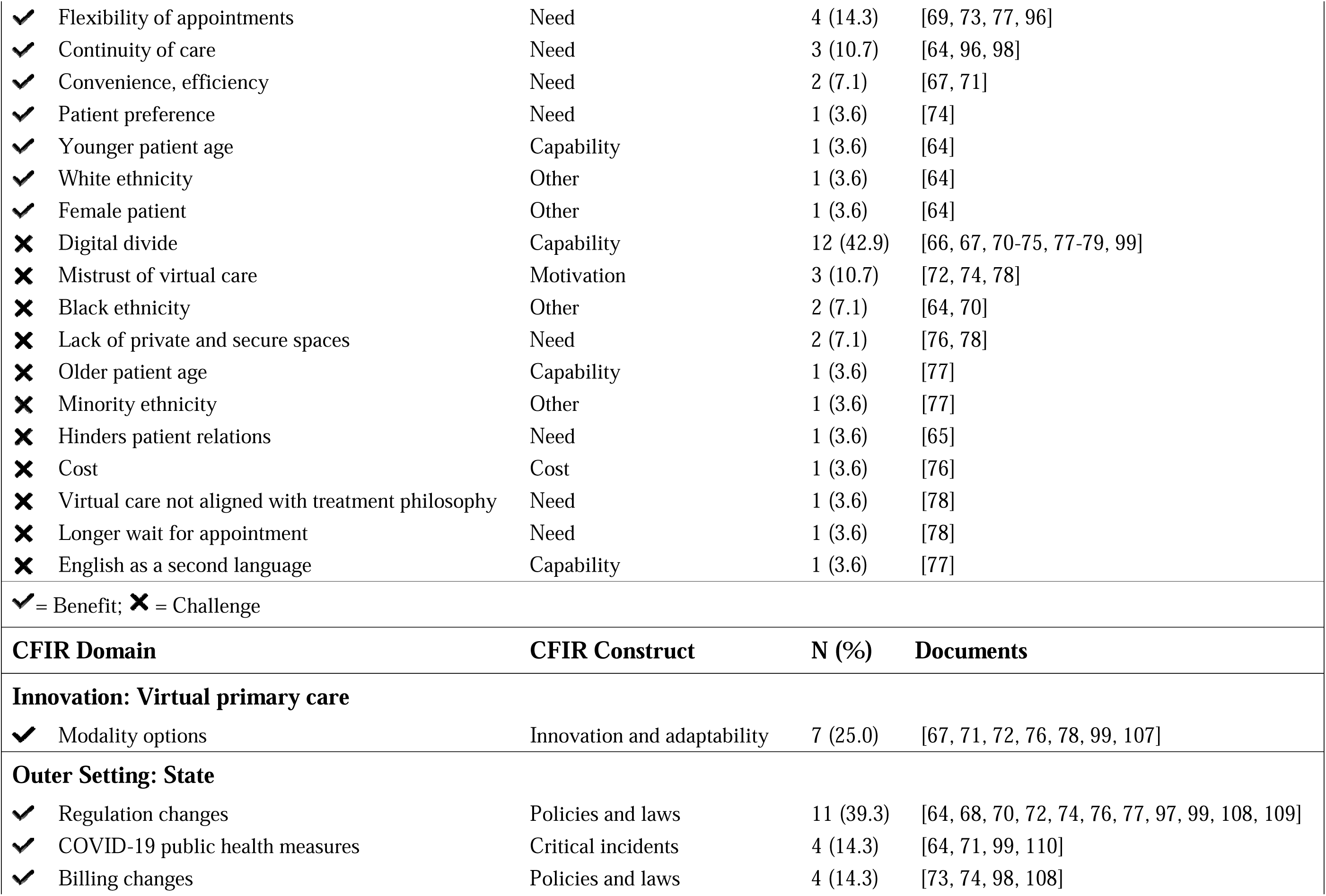
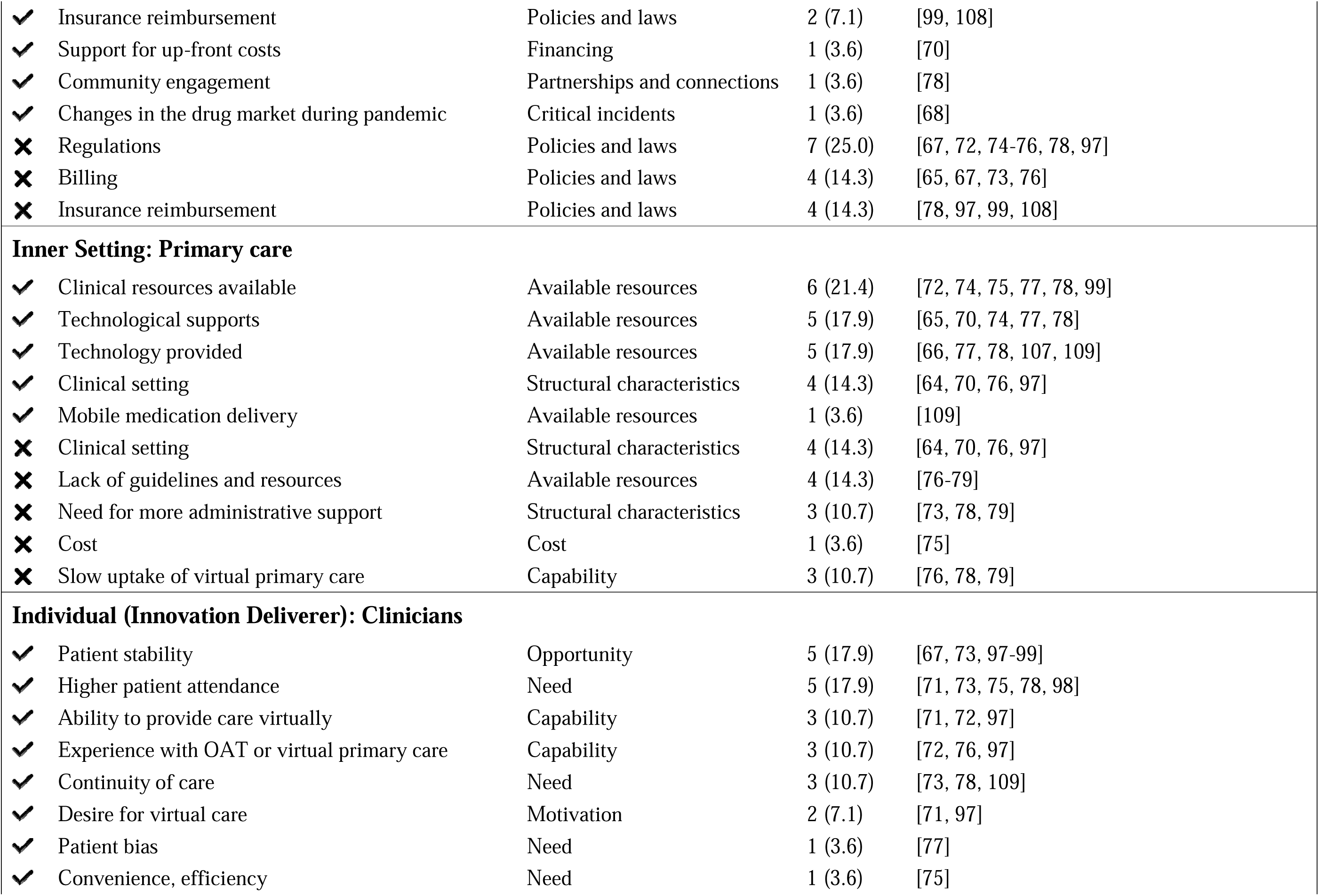
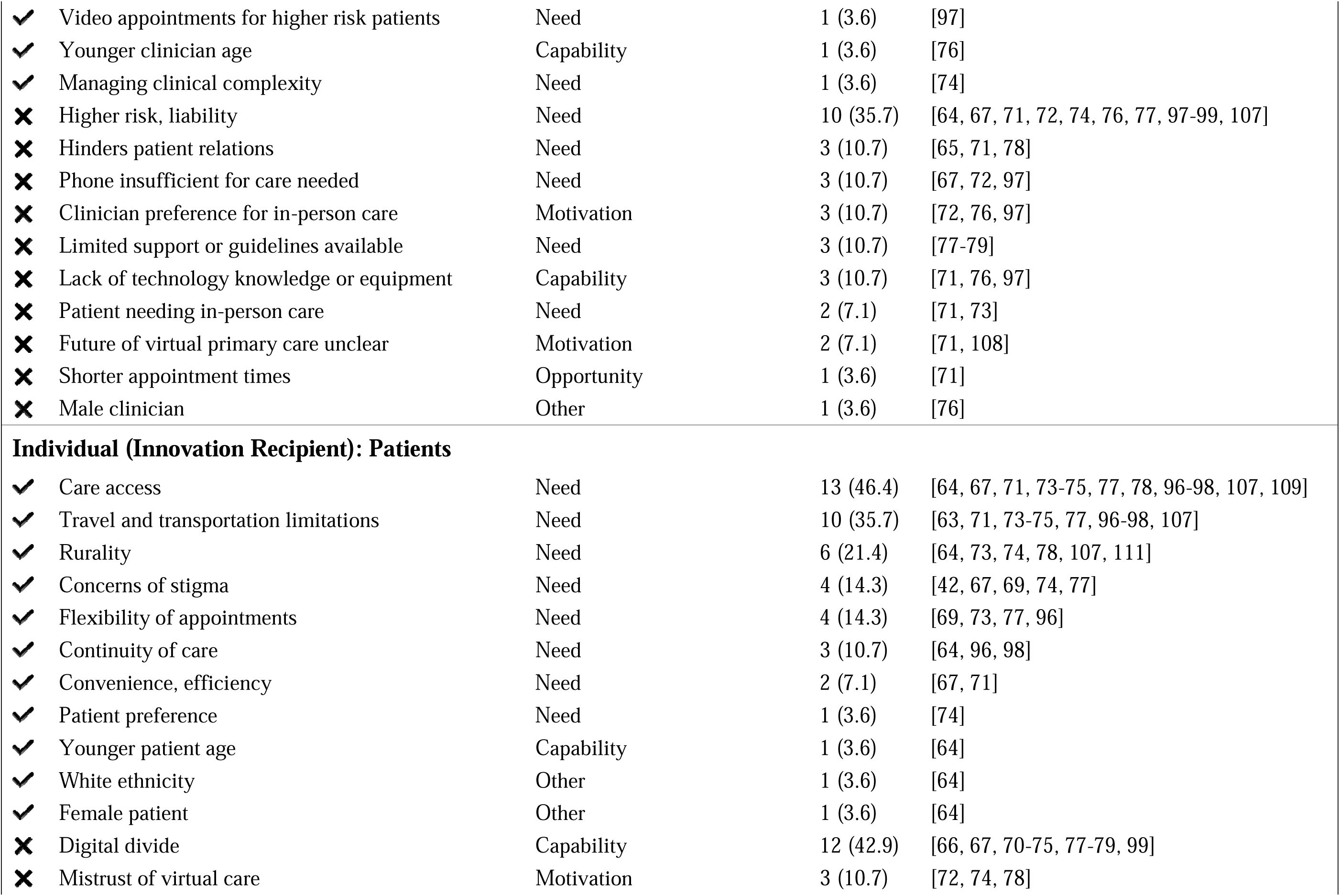
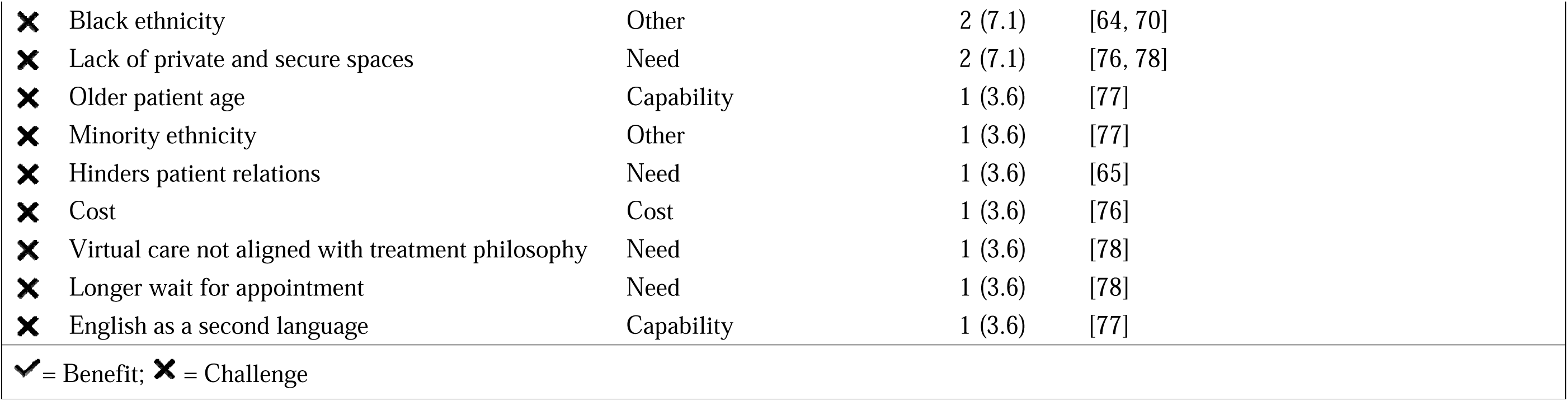
Benefits and challenges of virtual primary care for PWOUD by CFIR domains and constructs.

### Innovation: Virtual Care

Primary care (including but not limited to OAT initiation and management) was often delivered through telephone appointments (n=18, 64.3%), with fewer papers focusing on video modalities (n=14, 50.0%) or not specifying the type of synchronous virtual visits (n=9, 32.1%) (Table 4). Some studies emphasized the importance of providing multiple modality options such as virtual and in-person appointments (i.e., hybrid) (n=7, 25.0%).

**Table 4.**
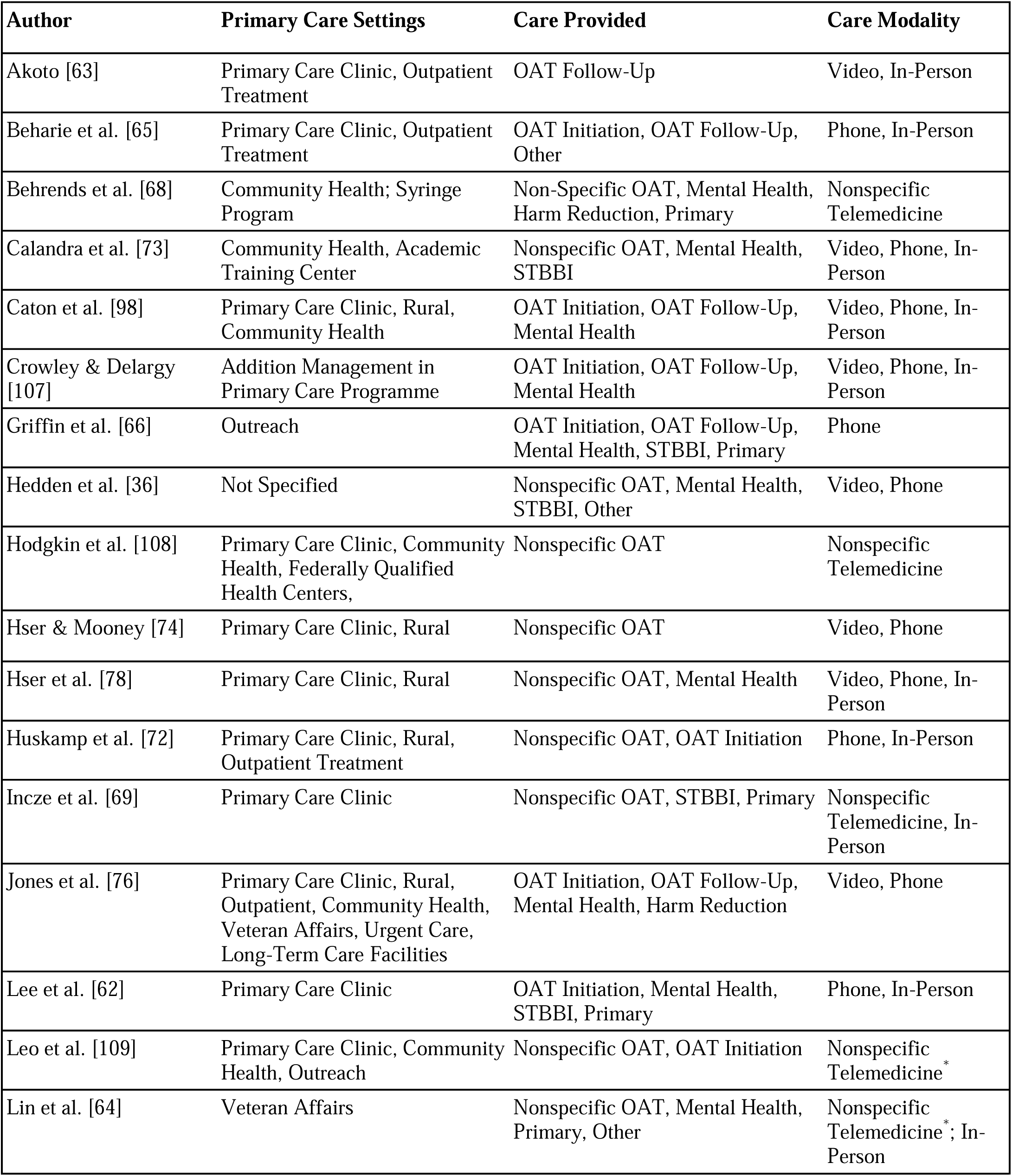
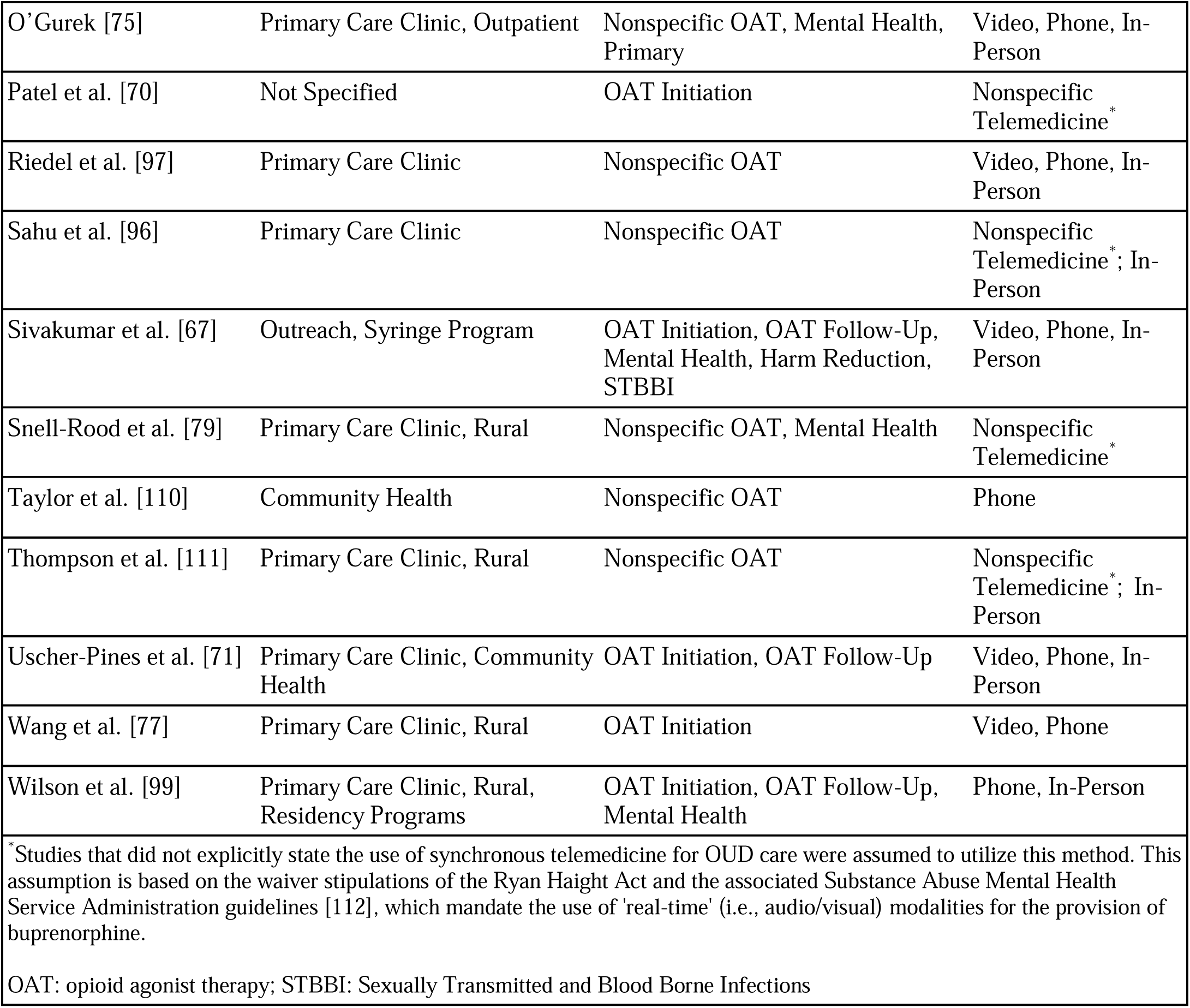
Primary care setting, care provided, and care modality.

Where studies described primary care prescription of OAT, most referred to provision generally, without specific mention of the stage or length of care (n=16, 57.1%). Others described OAT initiation (n=13, 46.4%) and/or follow-up and stabilization (n=9, 32.1%) for management of OUD. Outside of providing OAT, mental health care was the most commonly provided treatment (n=14, 50.0%), followed by treatments for sexually transmitted and blood-borne infections (n=6, 21.4%) [66]. Additionally, harm reduction services were provided (such as syringe exchange and naloxone distribution [67]), although to a lesser extent (n=3, 10.7%). Few studies (n=6, 21.4%) described other primary care needs (e.g., vaccinations [68], diabetes [69]), and lacked specific details on the care provided (e.g., was listed as ‘other health needs’ [65] or ‘other clinics’ [64]).

### Outer Setting: State

The most common facilitator described by included studies was regulatory changes (n=11, 39.3%) to support the delivery of virtual primary care for OUD, such as the waiver of the Ryan Haight Act in the United States which allowed clinicians to prescribe buprenorphine without the previously required first in-person appointment [70]. Another common facilitator was COVID-19 public health measures (n=4, 14.3%). Due to the increased COVID-19 risk to patients and clinicians associated with in-person appointments, PWOUD could utilize virtual primary care to adhere to social distancing measures and reduce their COVID-19 exposure risks [71].

Regulatory barriers were less commonly described (n=7, 25.0%) but included uncertainty in the permanence of the COVID-19 regulatory changes [72] and limited flexibility in providing methadone via virtual modalities [67]. Billing structures were described as both facilitators and challenges. For example, new billing structures or fee codes introduced during COVID-19 (n=5, 17.9%) facilitated virtual care delivery. However, some clinicians experienced challenges with billing (n=5, 17.9%), stating that payment models did not equally compensate virtual and in-person care [73] or that virtual engagement was not included as a billable service [65]. Modifications to insurance policies (n=4, 14.3%) yielded diverse perspectives with sufficient reimbursement for virtual services contingent upon policy provisions.

### Inner Setting: Primary Care

Several studies described clinical resources (n=6, 21.4%) as essential factors facilitating virtual primary care implementation, ensuring clinicians and patients could navigate virtual appointments. Such guidelines (n=6, 21.4%) ranged from documents produced by the Substance Abuse and Mental Health Services Administration to guide clinics through policy changes to providing OAT virtually [74] or individual clinic protocols for virtual delivery of OUD treatment [75]. Other resources included the provision of communication devices to clinicians and patients (n=5, 17.9%), and training or education for patients to prepare for their virtual appointments (n=5, 17.9%). Four studies (14.3%) noted that certain primary care clinics are better positioned to adopt virtual primary care. For example, clinics in urban or community-based settings [76] or larger clinics [70] are more likely to offer virtual OAT induction, although the reasoning behind these differences is not explained in the research.

Challenges affecting implementation at the clinic level included the need for more administrative support for physicians (n=4, 14.3%) and a lack of guidelines and resources (n=4, 14.3%) to support clinicians in providing evidence-based care (e.g., lack of protocols for virtual primary care [77]). Studies also highlighted the challenges associated with specific clinical settings (n=4, 14.3%), noting that accessing OUD care without an in-person examination was more difficult in solo-based primary care practices versus specialty substance use facilities, opioid treatment programs, community clinics, and veteran health administrations [76].

Furthermore, primary care practices in rural settings were less likely to provide virtual primary care services, possibly due to the lower demand for virtual services as COVID-19 risk was lower compared to urban areas [76]. A few studies described low demand for virtual care in general (n=3, 10.7%), both by patients and clinicians [76, 78, 79], resulting in slower uptake in some clinics.

### Individual Characteristics (Recipient): PWOUD

The most common benefit for patients was the high accessibility of virtual care (n=13, 46.4%), particularly for rural patients (n=6, 21.4%). This advantage was linked with other benefits, such as alleviating travel challenges (n=10, 35.7%) and flexibility of appointments (n=4, 14.3%). Factors like confidence in virtual care (n=2, 7.1%), its perceived convenience and efficiency (n=2, 7.1%), improved continuity of care (n=3, 10.7%), and the ability of virtual modalities to address patients’ stigma concerns (n=4, 14.3%) also emerged as pivotal factors. Additionally, a younger age demographic (n=1, 6.9%) and cost savings (n=2, 7.1%) positively influenced patients’ engagement with virtual primary care.

The gap between people who can easily use and access technology and those who cannot (i.e., “the digital divide”) remains the primary obstacle to the uptake of virtual care for PWOUD (n=12, 42.9%). This is compounded by a lack of technological knowledge or equipment (n=6, 21.4%), mistrust of virtual care (n=3, 10.7%), and absence of a private or secure space for appointments (n=2, 7.1%). Moreover, specific patient attributes intersected with virtual primary care adoption due to structural barriers of the healthcare system, such as being an older adult (n=1, 3.6%) and patients who experience racialization (n=3, 10.7%).

### Individual Characteristics (Provider): Clinicians

Clinicians identified several benefits of virtual care, particularly for patients who demonstrate stability (n=5, 17.9%) and can attend appointments consistently (n=5, 17.9%). Prioritizing continuity of care (n=3, 10.7%) also emerged as a strength. Furthermore, prior experience with OAT or using virtual modalities (n=3, 10.7%), and possessing a genuine interest in delivering virtual primary care (n=2, 7.1%) facilitated its adoption.

Conversely, apprehension about liability or heightened risks associated with offering virtual primary care for PWOUD (n=10, 35.7%) were seen as challenges among clinicians. Concerns also emerged over the adequacy of phone-based appointments relative to video (n=3, 10.7%), challenges of fostering a therapeutic relationship through virtual visits (n=3, 10.7%), and the belief that in-person care was necessary for patients (n=2, 7.1%). These sentiments resonated with clinicians’ preference for in-person care (n=3, 10.7%). Additional challenges included insufficient guidance for clinicians (n=3, 10.7%), such as evidence-based protocols [77], as well as limited technological understanding and inadequate equipment (n=3, 10.7%).

### Quality Appraisal

Over a quarter of the studies (n=8, 28.6%) received high scores through the QuADS tool, most of which were original research. In contrast, commentaries (n=5, 17.9%) and conference abstracts (n=5, 17.9%) predominantly received lower scores. Scores produced by our quality assessment using the QuADS tool are grouped by study type rather than an overall ranking (Table 5).

**Table 5.**
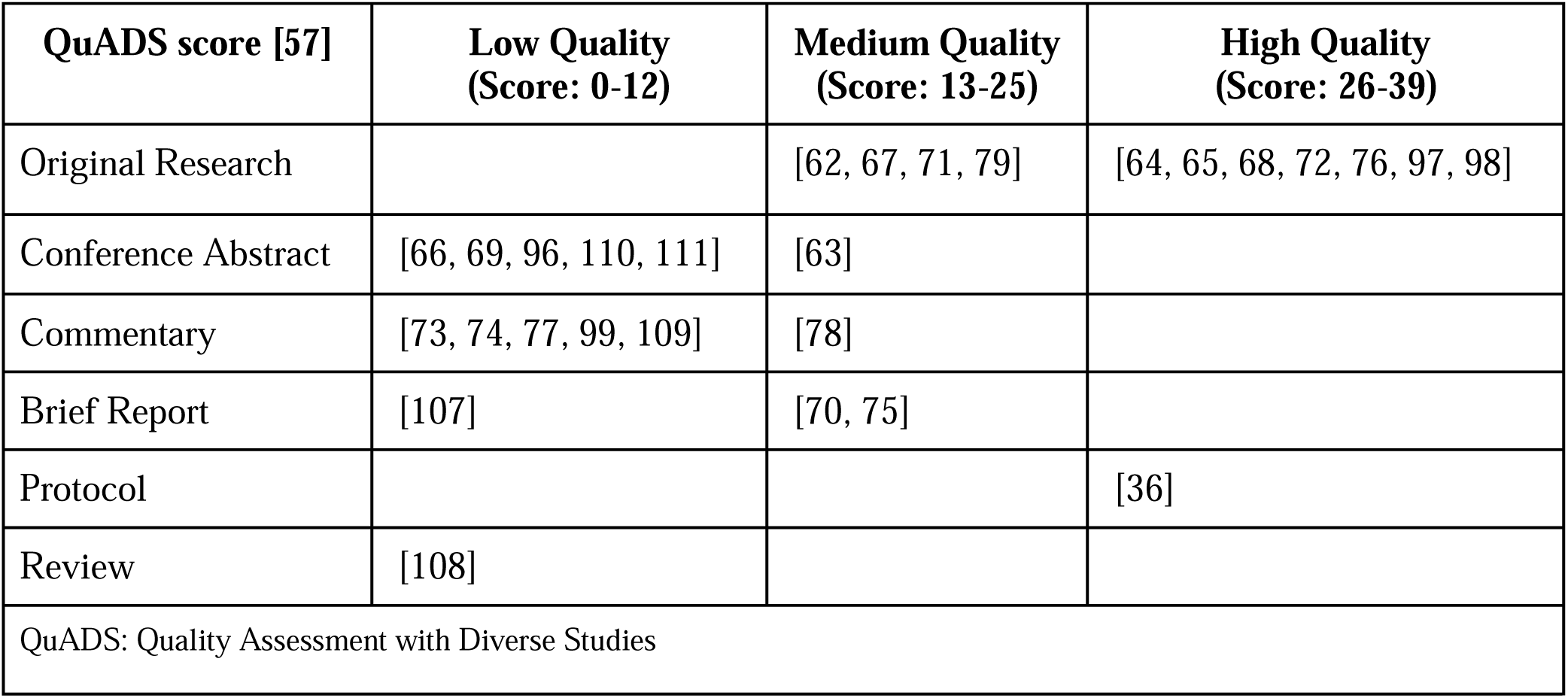
Quality appraisal.

## DISCUSSION

In reviewing 28 studies on the experiences of PWOUD and their primary care clinicians with virtual primary care, we sought to explore the current state of the literature, identify research gaps, and provide insights into the potential benefits and challenges of virtual approaches. The CFIR constructs were instrumental in our investigation, guiding us in identifying the organizational, structural, and individual-level factors that influence the implementation and ongoing use of virtual primary care for PWOUD. Our findings underscore that, with careful consideration of the implementation factors at all levels, virtual primary care holds promise as a feasible, acceptable, and effective option for PWOUD as complements to in-person primary care, aligning with research in other settings [80–82].

Notably, our review highlights that research related to virtual primary care for treating OUD is limited, and even more so for the broad range of primary care needs of PWOUD beyond OAT, despite the rapid increase in availability resulting from the COVID-19 pandemic. Further, despite the growing demand for research that distinguishes between experiences associated with sex and gender [83, 84], many of the studies we reviewed appear to conflate these two distinct experiences (e.g., reporting participant gender as female or male) or did not report any sex or gender-based analyses. Additionally, across the studies we reviewed, the heterogeneous research methods complicate data comparison and aggregation. Most studies were conducted in the United States, which can limit or challenge generalizability to other health system contexts. For example, regulations and billing, part of CFIR’s Outer Setting, differ dramatically across settings [85–87]. While qualified physicians in both countries can prescribe buprenorphine, the United States previously required a waiver under the Drug Addiction Treatment Act, whereas there is no such requirement in Canada [88]. Similarly, methadone maintenance treatment requires access to specialized clinics (i.e., opioid treatment programs) in the United States [89] but, in Canada, it can be prescribed in various healthcare settings [90].

PWOUD are known to frequently experience co-morbid conditions (e.g., HIV/AIDS, HCV, HBV [91]), infections (e.g., endocarditis, septic arthritis [92]), and other frequent/episodic healthcare needs (e.g., wound care [93]) that require timely and ongoing primary care. Notably, the extant literature focuses on treatment for OUD, and the studies we included did not routinely report how or if these conditions were recognized or treated. This remains a core knowledge gap. Future research on virtual primary care interventions for PWOUD would benefit from a more comprehensive description of the patients included and the range of care provided.

Relying on virtual modalities to increase access to healthcare was a common finding [94]. Virtual visits remove the necessity for physical travel to medical appointments and reduce associated costs (e.g., transit/gas, taking time off work, childcare). By offering the convenience of accessing care from a smartphone or other communications devices, virtual care can also support continuous access to and retention on OAT [95]. Patient lifestyle and stability play a significant role in driving their utilization of virtual care. Individuals with childcare responsibilities [77, 96], limited flexibility during work hours [73, 96], and those who are more stable on OAT [73, 97–99] may find virtual care well-suited to meeting their care needs, while minimizing disruption to their day-to-day lives.

The digital divide emerged as a central theme in multiple studies. Furthermore, clinicians’ belief that virtual primary care was unsuitable for some patients, deemed too risky due to patients’ health status, or associated with high liability also hindered its adoption, aligning with concerns found in other (i.e., non-primary care) clinical settings [100, 101]. With the challenges associated with patient stability and social determinants of health (e.g., digital technology access, longevity of OAT treatment, housing), providing care to PWOUD using solely virtual modalities may unintentionally limit equitable health care access. However, clinicians who believe virtual primary care is unsuitable for unstable patients may be ignoring patient preferences [74]. While virtual primary care for less stable patients is a nuanced topic and needs concerted research efforts and practice approaches, evidence is emerging that virtual primary care can provide a safe supplement to in-person visits for unstable or new patients [72, 97, 102]. To enhance the effectiveness of virtual primary care for this population, offering diverse appointment types can accommodate patient preferences, socio-economic factors, and health circumstances, fostering patient autonomy and promoting patient-centred care.

Virtual primary care for PWOUD is promising and feasible. However, this review also highlights that there may be disparities based on patient age [64, 77], language [77], race/ethnicity [64, 70, 77], sex [64], and those who experience the digital divide [66, 67, 70–75, 77–79, 99], due to rurality, socioeconomic status, or limited access to technology. These disparities pose challenges for implementing and accessing virtual care [103]. Without deliberate efforts to address these access barriers, virtual primary care could perpetuate inequities and worsen existing disparities in OAT access [104–106].

### Limitations

There are several limitations in this scoping review. Scoping reviews inherently possess limitations in terms of depth of analysis and generalizability [39], and the possibility of missing studies needs to be acknowledged. This is attributable to multiple factors; among them is our stringent inclusion criteria, which not only resulted in excluding studies with unclear or inadequately described settings but also affected our inter-rater reliability score. Furthermore, our definition of virtual visits excluded asynchronous forms, and we did not consider care settings providing only OAT. This criterion was in place to align with our broader research project [36] and to maintain a manageable review size. Additionally, studies published in languages other than English were not captured. There is also a variance in the quality of the included studies. Some included studies received low QuADS scores due to limitations in study information (i.e., conference abstracts with restricted word counts). Nonetheless, this scoping review presents novel results as the first of its kind covering virtual primary care for PWOUD. Our methodologically rigorous approach and the quality appraisal of the included studies make this review a valuable contribution to the literature, given the sparse research in this field.

### Conclusion

This scoping review highlights the promise of virtual primary care as a feasible, acceptable, and effective option for PWOUD, particularly in enhancing accessibility and continuous access to care. However, addressing the digital divide and mitigating disparities based on demographic and socioeconomic factors will be critical in realizing the full potential of virtual care. Future research and policy initiatives should prioritize efforts to bridge these gaps, ensuring that virtual primary care becomes an inclusive and equitable platform for delivering comprehensive care to and for PWOUD.

## Supporting information

PRISMA Abstract Checklist

PRISMA ScR Checklist

Data Extraction Template

## Data Availability

All data produced in the present work are contained in the manuscript

https://doi.org/10.1079/searchRxiv.2023.00329

https://doi.org/10.1079/searchRxiv.2023.00330

https://doi.org/10.1079/searchRxiv.2023.00331

https://doi.org/10.1079/searchRxiv.2023.00332

https://doi.org/10.1079/searchRxiv.2023.00333

## Abbreviations

OUD: Opioid Use Disorder
OAT: Opioid Agonist Treatment
PWOUD: People with Opioid Use Disorder
QuADS: Quality Assessment with Diverse Studies
CFIR: Consolidated Framework for Implementation Science Research
HIV/AIDS: Human Immunodeficiency Virus/Acquired Immune Deficiency Syndrome
HCV: Hepatitis C Virus
HBV: Hepatitis B Virus
STBBI: Sexually Transmitted and Blood Borne Infections

## Author contributions

Contributions to the paper are described using the CRediT taxonomy [113]. SN: Conceptualization, methodology, formal analysis, investigation, data curation, writing (original draft), writing (review and editing), visualization. EG: Conceptualization, methodology, formal analysis, investigation, data curation, writing (original draft), writing (review and editing), visualization. SS: Conceptualization, writing (review and editing), visualization. RM: Conceptualization, methodology, writing (review and editing), supervision, funding acquisition. LH: Conceptualization, methodology, writing (review and editing), supervision, funding acquisition. All authors have read and approved the final manuscript.

## Ethics approval and consent to participate

Not applicable.

## Consent for publication

Not required.

## Acknowledgements

We would like to thank the study team investigators for their valuable contributions and commitment to the project: Advancing Virtual Primary Care for People with Opioid Use Disorder.

