## Supplementary material for "Virtual Primary Care for People with Opioid Use Disorder: A Scoping Review of Current Strategies, Benefits, and Challenges": Data Extraction Template

**Supplementary Materials: Data Extraction Template**

**General information**

Title

Author details

Paper Type

- Original Research
- Brief Report
- Review Paper
- Conference Abstract
- Commentary
- Other

Country in which the study conducted

- United States
- UK
- Canada
- Australia
- Other

Publication date

Funding

**Characteristics of included studies**

Methods

Purpose of study

Study design

- Randomised controlled trial
- Non-randomised experimental study
- Cohort study
- Cross sectional study
- Case control study
- Systematic review
- Qualitative research
- Prevalence study
- Case series
- Case report
- Diagnostic test accuracy study
- Clinical prediction rule
- Economic evaluation
- Text and opinion
- Other

Population description

Total number of participants

Description of the intervention/Study Design

Type of care for PWOUD

- OAT initiation
- OAT follow up
- OAT/MOUD general (not specified)
- Syringe services
- STBBI/HCV/HIV
- Mental health (ex. Anxiety/Depression/PTSD)
- Other

Setting of primary care facility

- Family physician clinic
- Walk in clinic
- Rural clinic
- Veterans (VA Hospital/Clinic)
- Outpatient OAT/SUD treatment
- Community health centres/clinics
- Outreach/mobile clinic
- Primary care not specified
- Other

Modalities

- FaceTime/Zoom Call (Video Conference)
- Telephone (Audio only)
- In-person
- Telemedicine not specified
- Other

**Results**

Patient Outcomes or Results

Facilitators

Barriers

Relevant health system features

- Private fee and co-pay (finance)
- Free/universal access (finance)
- Medicare/Medicaid (finance)
- Uninsured (finance)
- Team-based care/interdisciplinary
- Clinicians/Teams Private (delivery)
- Clinicians/Teams Public (delivery)
- Low-barrier access (if yes include in other)
- Other

Future Research

Notes
